# Auditory and Visual Thalamocortical Connectivity Alterations in Unmedicated People with Schizophrenia: An Individualized Sensory Thalamic Localization and Resting-State Functional Connectivity Study

**DOI:** 10.1101/2024.12.18.24319241

**Authors:** John C. Williams, Philip N. Tubiolo, Roberto B. Gil, Zu Jie Zheng, Eilon B. Silver-Frankel, Natalka K. Haubold, Sameera K. Abeykoon, Dathy T. Pham, Najate Ojeil, Kelly Bobchin, Mark Slifstein, Jodi J. Weinstein, Greg Perlman, Guillermo Horga, Anissa Abi-Dargham, Jared X. Van Snellenberg

**Affiliations:** Department of Psychiatry and Behavioral Health, Renaissance School of Medicine at Stony Brook University, Stony Brook, NY 11794; Department of Biomedical Engineering, Stony Brook University, Stony Brook, NY 11794; Medical Scientist Training Program, Renaissance School of Medicine at Stony Brook University, Stony Brook, NY 11794; Scholars in BioMedical Sciences Training Program, Renaissance School of Medicine at Stony Brook University, Stony Brook, NY 11794; Department of Psychiatry, Columbia University Vagelos College of Physicians and Surgeons, New York-Presbyterian / Columbia University Irving Medical Center, New York, NY 10032; New York State Psychiatric Institute, New York, NY 10032; College of Medicine, State University of New York Downstate Health Sciences University, Brooklyn, NY 11203; Department of Neurobiology and Behavior, Cornell University, Ithaca, NY 14853; Department of Radiology, Renaissance School of Medicine at Stony Brook University, Stony Brook, NY 11794; Department of Psychology, Stony Brook University, Stony Brook, NY 11794

**Keywords:** Schizophrenia, thalamus, 22q11 deletion, sensory processing, thalamocortical, positive symptoms, medial geniculate nucleus, lateral geniculate nucleus, resting-state functional connectivity, functional localization, sensory thalamic localizer task, functional magnetic resonance imaging, auditory system, visual system

## Abstract

**Background:** Converging evidence from clinical neuroimaging and animal models has strongly implicated dysfunction of thalamocortical circuits in the pathophysiology of schizophrenia. Preclinical models of genetic risk for schizophrenia have shown reduced synaptic transmission from auditory thalamus to primary auditory cortex, which may represent a correlate of auditory disturbances such as hallucinations. Human neuroimaging studies, however, have found a generalized increase in resting state functional connectivity (RSFC) between whole thalamus and sensorimotor cortex in people with schizophrenia (PSZ). We aimed to more directly translate preclinical findings by specifically localizing auditory and visual thalamic nuclei in unmedicated PSZ and measuring RSFC to primary sensory cortices.

**Methods:** In this case-control study, 82 unmedicated PSZ and 55 matched healthy controls (HC) completed RSFC functional magnetic resonance imaging (fMRI). Auditory and visual thalamic nuclei were localized for 55 unmedicated PSZ and 46 HC who additionally completed a sensory thalamic nuclei localizer fMRI task (N = 101). Using localized nuclei as RSFC seeds we assessed group differences in auditory and visual thalamocortical connectivity and associations with positive symptom severity.

**Results:** Auditory thalamocortical connectivity was not significantly different between PSZ and HC, but hyperconnectivity was associated with greater positive symptom severity in bilateral superior temporal gyrus. Visual thalamocortical connectivity was significantly greater in PSZ relative to HC in secondary and higher-order visual cortex, but not predictive of positive symptom severity.

**Conclusion:** These results indicate that visual thalamocortical hyperconnectivity is a generalized marker of schizophrenia, while hyperconnectivity in auditory thalamocortical circuits relates more specifically to positive symptom severity.

## Introduction

Converging evidence from both clinical neuroimaging and animal models has strongly implicated dysfunction of thalamocortical circuits in the pathophysiology of schizophrenia (1–30). Notably, in a mouse model of the strongest known genetic risk factor for schizophrenia, 22q11.2 microdeletion syndrome (22q11DS) (31–38), synaptic transmission was found to be specifically impaired between the auditory thalamus (medial geniculate nucleus; MGN) and primary auditory cortex (AC; 19, 30). This impairment in MGN-AC connectivity, observed as weaker excitatory postsynaptic currents in AC following MGN stimulation, was evident in 22q11DS mice only after about 3 months of age (19), which is roughly equivalent to 20 years of age in humans. In comparison, the median age of onset of psychosis in individuals with 22q11DS is estimated to be 21 years (39), and symptoms of schizophrenia similarly typically emerge during late adolescence and early adulthood (40). Further, MGN-AC dysconnectivity in the 22q11DS mouse model was found to be caused by haploinsufficiency of the microRNA-processing gene Dgcr8, resulting in an excess of dopamine D2 receptors (D2Rs) in the thalamus and a hypersensitivity of MGN-AC projections to modulation by the antipsychotic medications haloperidol and clozapine (30). Finally, aberrantly reduced MGN-AC synaptic transmission was restored to healthy levels by administration of either haloperidol or clozapine, both of which attenuate positive symptoms of schizophrenia, including auditory hallucinations, in humans (19, 30). Positive symptoms are considered to be a core feature of the schizophrenia syndrome, and auditory hallucinations are estimated to be present in upwards of 70% of cases (41, 42), in addition to other auditory perceptual abnormalities (43). MGN-AC dysconnectivity may thus play a key role in the pathogenesis of schizophrenia generally, and potentially positive symptoms specifically, especially given the involvement of thalamic and cortical regions specific to auditory perception.

In humans, resting-state (RS) functional connectivity (RSFC) functional magnetic resonance imaging (fMRI) studies of schizophrenia and 22q11DS have identified widespread thalamocortical connectivity abnormalities, generally characterized by thalamic hyperconnectivity to sensorimotor cortical regions and hypoconnectivity to prefrontal cortex (PFC), as well as alterations in the volumes of various thalamic nuclei (1–16). In contrast to the hypoconnectivity that might be expected from the weaker synaptic transmission observed in the 22q11DS mouse model, these findings from human studies include thalamic *hyper*connectivity with AC (and visual cortex; VC), which could represent correlates either of schizophrenia generally or of positive symptom severity more specifically. One potential explanation for this discrepancy is that prior studies of thalamocortical connectivity in schizophrenia have not separately identified thalamic sensory nuclei, such as the MGN or lateral geniculate nucleus (LGN), and assessed their connectivity to sensory cortices independently from the rest of the thalamus. This is likely due to the difficulty in localizing MGN and LGN using standard T1-weighted anatomical imaging sequences (13), which would be needed to create MGN or LGN regions of interest (ROIs) for functional connectivity analyses.

Here, we sought replicate findings from the 22q11DS mouse in a study of human individuals with schizophrenia, reasoning that if the weaker excitatory postsynaptic currents observed in 22q11DS mice were present in people with schizophrenia (PSZ), these individuals should exhibit weaker RSFC between MGN and AC. In doing so, it was critical to employ a sample of unmedicated PSZ, because, as discussed above, antipsychotic medications have been shown to normalize MGN-AC synaptic transmission in 22q11DS mice (30). Consequently, we conducted a multi-modal fMRI study of unmedicated people with schizophrenia and matched healthy control participants (HC) who completed both RS fMRI and a sensory thalamic localizer (TL) fMRI task paradigm that we recently developed and validated (44) to allow participant-specific localization of the human auditory thalamus (MGN) and visual thalamus (LGN). We previously showed that the functionally defined ROIs (fROIs) obtained from this task outperform several atlas-based and automated segmentation-based methods for obtaining MGN and LGN ROIs (44), indicating that fROIs from the TL task should produce more accurate timeseries of BOLD signal in the MGN and LGN than alternative approaches to obtaining ROIs for these thalamic nuclei.

We used participant-specific MGN fROIs to investigate whether PSZ exhibit reduced MGN-AC connectivity relative to HC, and whether MGN-AC connectivity is associated with positive symptom severity, as measured using the Positive and Negative Syndrome Scale (PANSS; 45, 46). We then repeated these analyses using LGN fROIs that are also available through the TL task, to determine whether these findings are specific to auditory thalamocortical circuits as in the 22q11DS mouse model (30), or represent more generalized deficits in thalamocortical sensory processing pathways, given that visual hallucinations are not uncommon in schizophrenia, despite the predominance of auditory hallucinations (41, 42, 47). We additionally performed whole-cortex RSFC analyses using whole thalamus seeds obtained from the FreeSurfer thalamic segmentation (48), to help contextualize the present study with respect to prior studies of whole thalamus connectivity in schizophrenia.

## Materials and Methods

### Overview

This is a case-control study of unmedicated PSZ and HC demographically matched on age, sex, and parental SES, ages 18-60 years old, acquired across two sites: the New York State Psychiatric Institute (NYSPI) and Stony Brook University (SBU). All procedures at each institution were approved by their respective Institutional Review Board. Full inclusion and exclusion criteria for each site, as well as the demographic and clinical assessments employed, are described in the **Supplementary Materials and Methods**

### fMRI Acquisition and Analysis

MRI was completed at NYSPI using a 3T General Electric (Boston, MA) MR750 with a NOVA 32-channel head coil, and at SBU using a 3T Siemens (Munich, Germany) MAGNETOM Prisma at the SBU Social, Cognitive, and Affective Neuroscience (SCAN) Center with a Siemens 64-channel head-and-neck coil (49). Participants were asked to complete 4 T2* BOLD fMRI runs each of RS fMRI and the TL fMRI task, with a minimum of two runs of each required for inclusion in the final analyzed dataset. BOLD fMRI runs were acquired in alternating anterior-posterior and posterior-anterior phase-encode directions. The sensory TL task, based on prior work (50, 51), enables the acquisition of participant-level functionally defined regions of interest (fROIs) for the MGN (auditory thalamus) and LGN (visual thalamus), by identifying thalamic regions specifically coactivating with AC and visual cortex (VC), respectively, during alternating periods of auditory and visual stimulation. This task and analysis pipeline used to generate thalamic fROIs is freely available and has been described in detail and validated previously (44); a summary is provided in the ***fMRI Task Procedures*** section of **Supplementary Materials and Methods.**

Each RS run was 7 min 34 s at SBU, and 7 min 38 s at NYSPI; each TL run was 3 min 46 s at both sites. The following were also acquired in each MRI session: 1) high-resolution T1-weighted and T2-weighted anatomical scans with 0.80mm isotropic voxels, 2) spin echo field maps in the anterior-posterior and posterior-anterior directions, and 3) a B0 field map. BOLD fMRI data were acquired at NYSPI with a repetition of 850 ms, and field-of-view of 192 mm, and at SBU with a repetition time of 800 ms and field-of-view of 204 mm. At both sites, data were acquired with an echo time of 25 ms, multiband factor of 6, flip angle of 60 degrees, and 2 mm isotropic voxels.

RS and TL fMRI data were preprocessed using the Human Connectome Project Minimal Preprocessing Pipelines, and were then processed through a standardized pipeline, described in full in the *fMRI Pre- and Post-Processing* section of Supplementary Materials and Methods.

### Resting-State Functional Connectivity Analyses

#### Regions of Interest

Medial geniculate nucleus (MGN) and lateral geniculate nucleus (LGN) fROIs were obtained for participants with at least 2 usable runs each of RS data and the sensory TL task. ROIs for mediodorsal nucleus (MD) and whole thalamus were each obtained from the FreeSurfer thalamic nuclei segmentation (48) for each participant with 2 usable RS runs using their participant’s anterior commissure-posterior commissure aligned, readout distortion and bias field corrected T1-weighted image. The MD ROI was generated by uniting the medial magnocellular and lateral parvocellular segments, and the thalamus ROI was generated by uniting all obtained thalamic segments. Left and right MGN, LGN, and thalamus time series were extracted from each RS run’s post-processed NIfTI image for use as RSFC seeds.

An AC mask for group-level analyses was generated using the Glasser parcellation (52), by selecting all parcels within the “auditory” network of the Cole-Anticevic Brain Network Partition (53) and dilating by 4 mm. A VC mask was produced similarly, but by instead selecting parcels within the “visual1” and “visual2” networks (as primary and secondary visual regions are in separate partitions), and retaining only the largest contiguous cluster (in order to remove small visual parcels that are discontinuous with the main visual occipital region in this partition), before dilating by 4mm.

#### Seed Resting-State Functional Connectivity

Whole-cortex seed RSFC images were generated for each seed in each hemisphere (left and right MGN, LGN, and whole thalamus) by calculating the Pearson’s partial correlation between each run’s CIFTI grayordinate time series and extracted seed time series. Partial correlations included nuisance regressors for WM, CSF, and global signal, and their derivatives, as well as the 6 fMPs and their derivatives, squares, and squared derivatives. Left and right seed RSFC images were averaged to produce a single run-level seed RSFC image for each seed, which were then averaged across available RS runs to create a participant-level seed RSFC image for each participant.

To evaluate the specificity of MGN and LGN seed connectivity associations detected in subsequent analyses, which are described in the ***Post-Hoc Specificity Analyses*** section of **Supplementary Materials and Methods**, additional MGN and LGN seed RSFC analyses were conducted using thalamic signals as additional nuisance regressors when calculating partial correlations (i.e., in addition to the nuisance regressors already used), as follows: 1) MGN RSFC images were generated while controlling for LGN and MD timeseries, and 2) LGN RSFC images were generated while additionally controlling for MGN and MD timeseries.

CIFTI seed correlation images were harmonized across sites using ComBat (54–57). Covariates included during ComBat harmonization were age, biological sex, handedness, race, ethnicity, and parental SES, diagnosis (PSZ or HC), and antipsychotic medication exposure (greater than 2 weeks of cumulative lifetime exposure to antipsychotic medications), as well as the positive, negative, and general subscales from the PANSS. Imputed values were used in place of any missing data demographic or PANSS assessment data (see ***Multiple Imputation*** in **Supplementary Materials and Methods**). CIFTI seed RSFC images were then separated into left and right cortical surface seed RSFC GIFTI images using Connectome Workbench (58) for group-level analyses.

### Statistical Analysis

Statistical analyses were performed in MATLAB R2018a (The MathWorks, Inc., Natick, MA). Effect sizes for group differences in demographics were calculated using Hedges’s g (59) or Cramér’s V (60), as appropriate. Group differences in PANSS scores were assessed for significance using Mann-Whitney-Wilcoxon rank-sum tests (61, 62).

Group-level localization analyses were performed in Permutation Analysis of Linear Models (PALM) (63–67), using 10,000 permutations or sign-flips, family-wise error rate corrected (68) over tested grayordinates, threshold-free cluster enhancement (TFCE; 69), and mean-centering of data and design matrix columns except where otherwise specified. Age, biological sex, and handedness were included as covariates.

Group differences in seed connectivity were assessed using a model that included a dichotomous schizophrenia diagnosis regressor and covariates; associations with symptom severity were assessed using a model including each of the PANSS subscales (PS, NS, and GS) and schizophrenia diagnosis and covariates. Cortical regions exhibiting significant seed RSFC in each group (HC and PSZ) were determined using a model consisting of an intercept only, without mean-centering data. T-statistic maps were thresholded at α *=* 0.05 after Bonferroni correction over hemispheres.

Primary analyses assessing the significance of group differences and associations with positive symptoms in MGN and LGN fROI seeds were conducted within AC and VC masks (described in ***Regions of Interest,*** above) and small-volume corrected (68). Whole-cortex follow-up analyses were then conducted to identify regions showing significant LGN and MGN seed connectivity in each group (PSZ and HC), as well as the difference in seed connectivity between groups. Analyses using whole thalamus seeds were conducted in whole cortex only.

Missing demographic data were estimated using multiple imputation for use as covariates (see ***Multiple Imputation*** in **Supplementary Materials and Methods**); participants with missing PANSS assessment data were excluded from analysis testing symptom associations. To visualize identified associations, seed RSFC from grayordinates displaying significant associations were additionally extracted and used as dependent variables for generalized linear models using the same model used to identify them in PALM, using Huber robust regression (70).

## Results

### Participants

Eighty-two PSZ and 55 matched HC completed at least 2 runs of RS fMRI (RS-only; N = 137), with PANSS assessments available for 78 PSZ and 51 HC (N = 129), after excluding 2 PSZ and 1 HC for excess motion after volume censoring. Sixty-two PSZ and 50 HC additionally completed at least 2 runs of TL task fMRI; after quality checks, TL fROIs were excluded for 7 PSZ and 4 HC, producing a final dataset of 55 PSZ and 46 HC after volume censoring (RS+TL; N = 101); PANSS assessments were additionally available for 52 PSZ and 42 HC (N = 94). Analyses using TL MGN and LGN fROI seeds used the RS+TL sample, while analyses using whole thalamus seeds used the larger RS only sample to maximize power.

Summary statistics for demographics and clinical assessments for the RS-only and RS+TL case-control samples are shown in **Table 1**. PANSS symptom severity, visualized in **Figure S1,** is broadly similar to past thalamocortical RSFC studies schizophrenia that utilized samples that included predominately medicated PSZ samples (e.g., 6, 7, 8, 10).

**Table 1.** Participant Demographics and Clinical Measures.

| Participant Characteristic | RS-only (N = 137) |  |  | RS+TL (N = 101) |  |  |
| --- | --- | --- | --- | --- | --- | --- |
|  | PSZ (n = 82) | HC (n = 55) | Effect Size<sup>a</sup> | PSZ (n = 55) | HC (n = 46) | Effect Size<sup>a</sup> |
| Age, Years (SD) | 31.9 (11.9) | 30.7 (10.2) | -0.106 | 32.2 (11.8) | 30.0 (9.9) | -0.206 |
| Biological Sex, Female (%) | 30 (36.6%) | 18.0 (32.7%) | 0.040 | 18.0 (32.7%) | 14.0 (30.4%) | 0.025 |
| Ethnicity, Hispanic (%) | 20.0 (24.4%) | 15.0 (27.3%) | 0.197 | 13.0 (23.6%) | 14.0 (30.4%) | 0.126 |
| Race |  |  | 0.200 |  |  | 0.267 |
| African American (%) | 28.0 (34.1%) | 22.0 (40.0%) |  | 19.0 (34.5%) | 17.0 (37.0%) |  |
| Asian (%) | 6.0 (7.3%) | 5.0 (9.1%) |  | 3.0 (5.5%) | 5.0 (10.9%) |  |
| Caucasian (%) | 28.0 (34.1%) | 19.0 (34.5%) |  | 18.0 (32.7%) | 15.0 (32.6%) |  |
| More than one (%) | 13.0 (15.9%) | 2.0 (3.6%) |  | 11.0 (20.0%) | 2.0 (4.3%) |  |
| Other (%) | 7.0 (8.5%) | 7.0 (12.7%) |  | 4.0 (7.3%) | 7.0 (15.2%) |  |
| Handedness<sup>b</sup> |  |  | 0.122 |  |  | 0.122 |
| Ambidextrous (%) | 9.0 (11.0%) | 3.0 (5.5%) |  | 6.0 (10.9%) | 2.0 (4.3%) |  |
| Left (%) | 2.0 (2.4%) | 3.0 (5.5%) |  | 2.0 (3.6%) | 2.0 (4.3%) |  |
| Right (%) | 71.0 (86.6%) | 49.0 (89.1%) |  | 47.0 (85.5%) | 42.0 (91.3%) |  |
| Smoker<sup>c,d</sup> (%) | 28.0 (34.1%) | 3.0 (5.5%) | -0.440 | 19.0 (35.2%) | 3.0 (6.5%) | -0.522 |
| Parental SES<sup>e,f</sup> (SD) | 38.2 (14.3) | 44.0 (10.7) | 0.348 | 41.0 (13.4) | 43.9 (11.5) | 0.228 |
| PANSS Total<sup>g</sup> (SD) | 69.9 (17.5) | 34.9 (6.3) | -2.459 | 70.0 (17.1) | 35.0 (6.7) | -2.563 |
| PANSS General<sup>g</sup> (SD) | 35.9 (9.6) | 18.3 (3.6) | -2.257 | 35.9 (9.7) | 18.4 (3.8) | -2.276 |
| PANSS Negative<sup>g</sup> (SD) | 15.5 (6.0) | 9.2 (3.1) | -1.249 | 15.5 (5.4) | 9.2 (3.2) | -1.362 |
| PANSS Positive<sup>g</sup> (SD) | 18.5 (6.1) | 7.4 (0.9) | -2.317 | 18.6 (6.5) | 7.4 (0.9) | -2.275 |
| PSYRATS Total<sup>h</sup> (SD) | 25.0 (19.7) | 2.1 (10.6) | -1.279 | 24.3 (19.2) | 2.5 (11.5) | -1.226 |
| PSYRATS AH<sup>i</sup> (SD) | 12.8 (14.8) | 1.3 (6.9) | -0.853 | 11.8 (14.4) | 1.6 (7.5) | -0.796 |
| PSYRATS D<sup>h</sup> (SD) | 11.5 (7.8) | 0.7 (3.7) | -1.530 | 11.0 (7.4) | 0.9 (7.5) | -1.423 |
| SANS<sup>j</sup> (SD) | 7.3 (5.8) | 2.0 (3.2) | -0.998 | 9.6 (4.5) | 2.2 (3.4) | -1.729 |
| CDRS<sup>k</sup> (SD) | 4.4 (4.4) | 0.3 (1.0) | -1.1414 | 4.7 (4.6) | 0.3 (1.0) | -1.199 |
| AP Medication-Naïve<sup>l</sup> (%) | 31 (37.8%) | — | — | 19 (34.6%) | — | — |

<sup>a</sup>Effect sizes for continuous variables were calculated with Hedges's g; effect sizes for dichotomous variables were calculated with Cramér's V.
<sup>b</sup>Handedness was reported using the Edinburgh Handedness Inventory.
<sup>c</sup>Smoking status defined as current daily nicotine use.
<sup>d</sup>Smoking status was unavailable for 2 participants (1 PSZ and 1 HC) from the RS-only sample and 1 PSZ from the RS+TL sample.
<sup>e</sup>Mean Parental SES was reported using the Hollingshead Four Factor Index Scale of Socioeconomic Status scale.
<sup>f</sup>Parental SES data were unavailable for 13 participants (10 PSZ and 3 HC) from the RS-only sample and 9 participants (6 PSZ and 3 HC) from the RS+TL sample.
<sup>g</sup>PANSS data were unavailable for 8 participants (4 PSZ and 4 HC) from the RS-only sample and 7 participants (3 PSZ and 4 HC) from the RS+TL sample.
<sup>h</sup>PSYRATS Total and PSYRATS D data were unavailable for 42 participants (13 PSZ and 29 HC) in the RS-only sample and 34 participants (10 PSZ and 24 HC) in the RS+TL sample.
<sup>i</sup>PSYRATS AH data were unavailable for 36 participants (7 PSZ and 29 HC) in the RS-only sample and 28 participants (4 PSZ and 24 HC) in the RS+TL sample.
<sup>j</sup>SANS data were unavailable for 40 participants (11 PSZ and 29 HC) in the RS-only sample and 31 participants (7 PSZ and 24 HC) in the RS+TL sample.
<sup>k</sup>CDRS data were unavailable for 22 participants (10 PSZ and 12 HC) in the RS-only sample and 15 participants (5 PSZ and 10 HC) in the RL+TL sample.
<sup>l</sup>All PSZ in this study were antipsychotic medication-free, defined as no exposure to oral antipsychotic medication for at least 3 weeks, and no exposure to intramuscular antipsychotic medications for at least 6 months, prior to participation in the study. PSZ were additionally considered to be antipsychotic medication-naïve if they had fewer than 2 weeks of cumulative lifetime exposure to antipsychotic medications.
**Abbreviations:** PSZ: People with Schizophrenia; HC: Healthy Controls; SD: standard deviation; SES: socioeconomic status; PANSS: Positive and Negative Syndrome Scale; PSYRATS: Psychotic Symptom Rating Scales; PSYRATS AH: PSYRATS Auditory Hallucinations; PSYRATS D: PSYRATS Delusions; SANS: Scales for the Assessment of Negative Symptoms; CDRS: Calgary Depression Scale for Schizophrenia. AP: Antipsychotic.

### Functionally Defined Regions of Interest

Probability density images for the MGN and LGN fROIs obtained in the RS+TL sample are shown **Figure S2**, and for whole thalamus ROIs obtained in the RS-only sample in **Figure S3.** AC and VC masks used for TL fROI seed correlation group-level analyses in PALM are shown in **Figure S4.**

### MGN-AC and LGN-VC Seed Connectivity

#### Positive Symptom Associations

Positive symptom severity was significantly associated with greater MGN-AC RSFC (hyperconnectivity) in a region of association AC on the superior temporal gyrus (STG) **(Figure 1A-C).** Post-hoc analyses indicated that this relationship is specific to connectivity from MGN (and not LGN or MD; **Table S1**) and is robust to controlling for signal from LGN and MD **(Table S2),** although doing so does produce a small but statistically significant reduction in the observed relationship **(Table S3).** No significant associations were found between LGN-VC RSFC and positive symptom severity.

**Figure 1.**
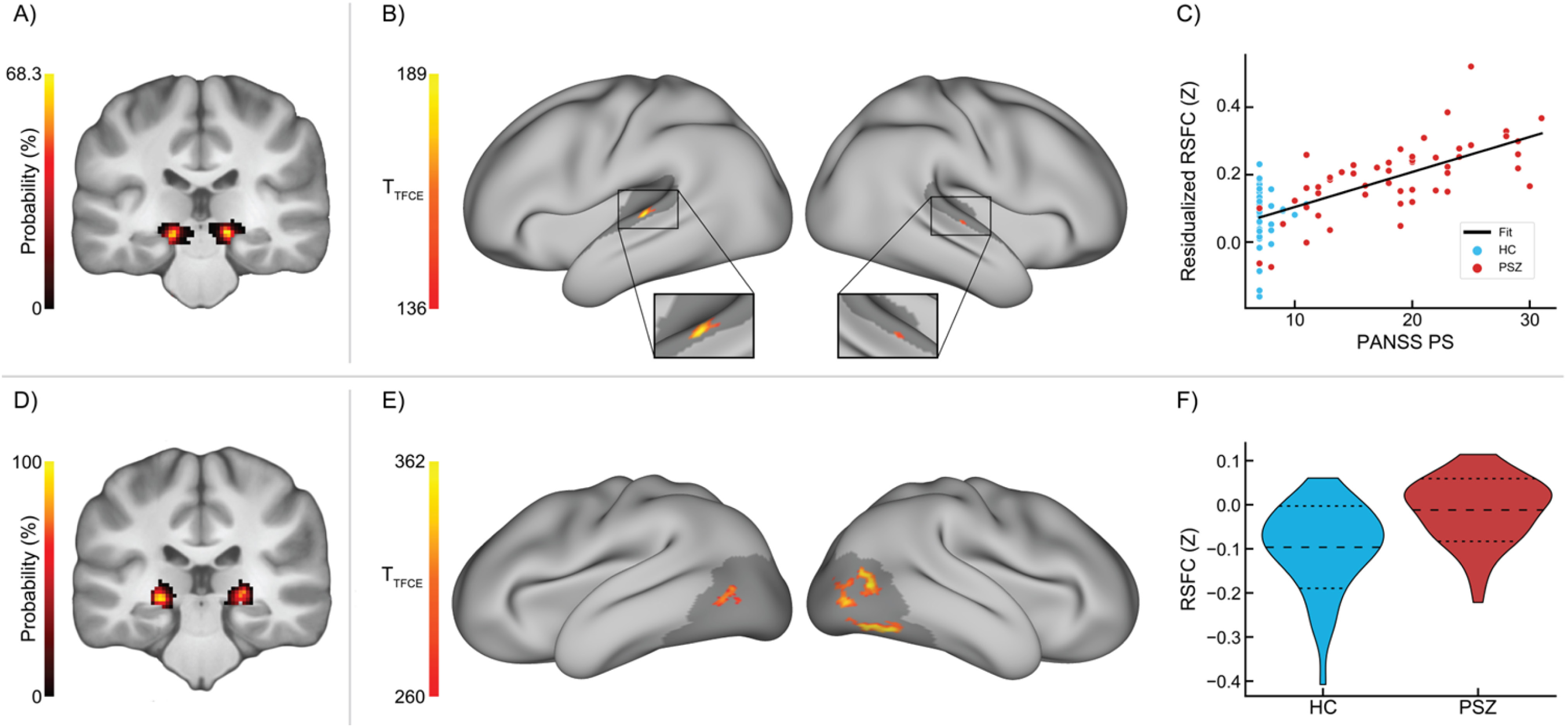
T-statistic maps for thalamic localizer (TL) functionally defined regions of interest (fROIs) for group differences and associations with positive symptom severity in medial geniculate nucleus (MGN) and lateral geniculate nucleus (LGN) seed resting-state functional connectivity (RSFC) in the RS+TL sample (total N = 101). Significance was assessed grayordinate-wise using cortical surface GIFTIs in Permutation Analysis of Linear Models (PALM) within auditory cortex (AC) and visual cortex (VC) masks, using 10,000 permutations and threshold-free cluster enhancement. P-values were corrected for family-wise error rate; t-statistic maps were thresholded at α = 0.05. **A)** Probability density maps of MGN fROIs obtained from the TL task across participants in the RS+TL sample, in Montreal Neurological Institute 152 non-linear 6th-generation (MNI152NLin6) space. **B)** MGN RS seed connectivity associations with Positive and Negative Symptom Scale (PANSS) Positive subscale scores in auditory cortex (AC), controlling for age, biological sex, handedness, and schizophrenia diagnosis, as well as PANSS negative and general subscores. Gray underlay shows the auditory cortex mask used in PALM. 28 significant grayordinates. PANSS data were available for 52 patients with schizophrenia (PSZ) and 42 healthy controls (HC); total N = 94. **C)** MGN seed connectivity extracted from the significant grayordinates visualized in **B**, with MGN resting-state functional connectivity (RSFC) residualized to covariates, displaying the identified relationship between MGN seed connectivity and positive symptom severity. **D)** Probability density maps of LGN fROIs obtained from the TL task across participants in the RS+TL sample, in MNI152NLin6 space. **E)** LGN RS seed connectivity group differences between PSZ (n = 55) and HC (n = 46) in visual cortex (VC), controlling for age, biological sex, and handedness. 245 significant grayordinates. **F)** LGN seed connectivity extracted from the significant grayordinates visualized in **E**, displaying the distribution of LGN seed connectivity in HC and PSZ. Dashed lines in panels **C** and **F** indicate group means ± 1 standard deviation.

#### Group Differences

LGN-VC RSFC was significantly greater in PSZ relative to HC **(Figure 1D-F)** in right secondary and bilateral tertiary/higher-order VC, including right fusiform face area. Post-hoc analyses indicated that this relationship is specific to connectivity from LGN (and not MGN or MD; **Table S4**) and is robust to controlling for signal from MGN and MD **(Table S5),** although doing so does produce a small but statistically significant reduction in the observed relationship **(Table S6).** We found no significant group differences between PSZ and HC in MGN RSFC to any grayordinates in our AC ROI.

### MGN and LGN Whole-Cortex Seed Connectivity

Regions of significant MGN and LGN seed connectivity in PSZ and HC are shown for HC and PSZ in **Figure 2A-D**. Of note, MGN was significantly connected to primary and secondary AC in both groups, as well as posterior cingulate and retrosplenial cortex, medial PFC, parahippocampal area (including entorhinal cortex; PHA). Anticorrelations were observed with large areas of secondary and higher-order VC, posterior parietal cortex, lateral PFC, premotor cortex, motor cortex, and somatomotor cortex. In HC, anticorrelations were also observed in association AC.

**Figure 2.**
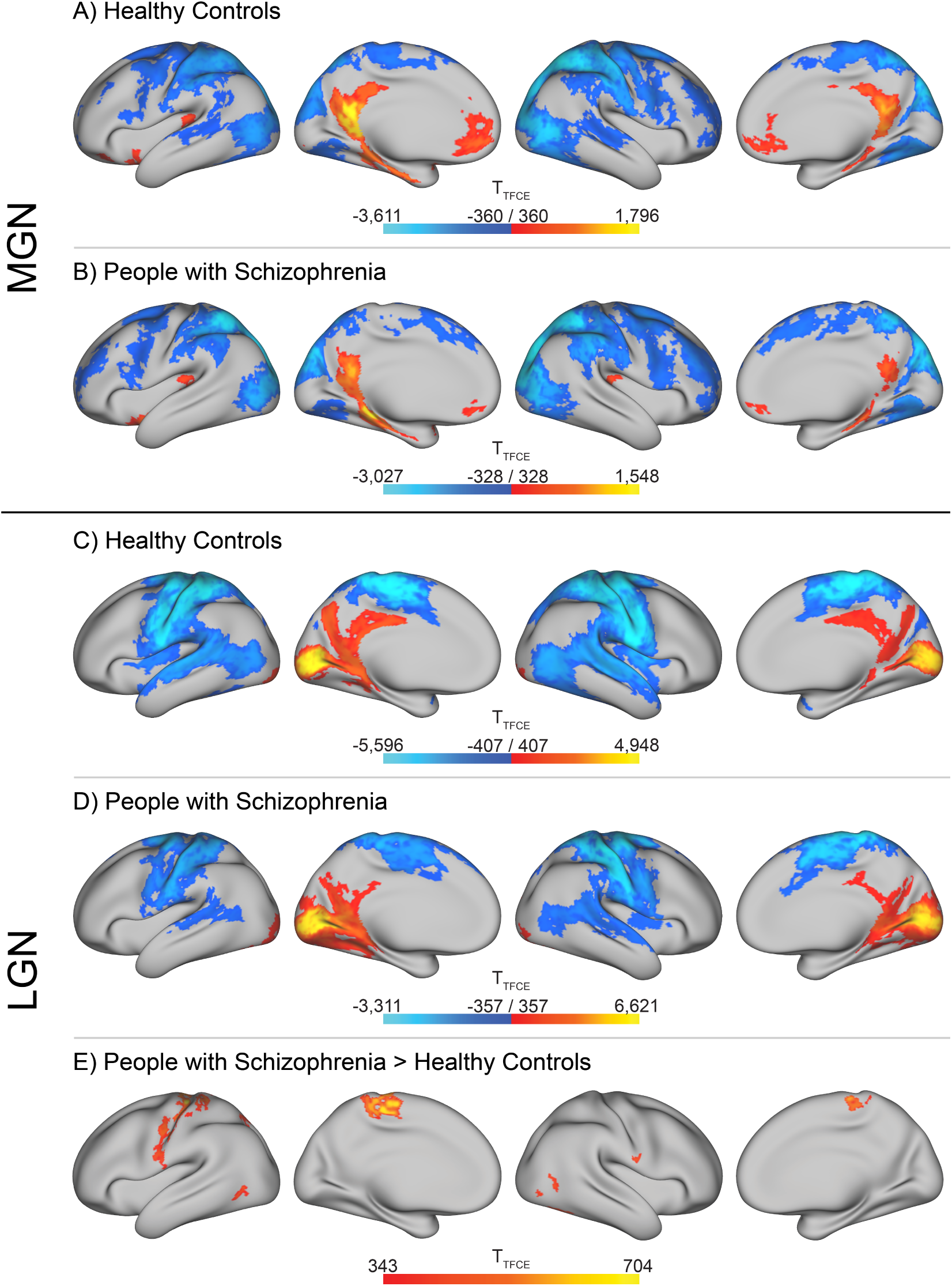
Panels **A-D** show whole-cortex t-statistic maps for grayordinates exhibiting significant resting-state functional connectivity (RSFC) from medial geniculate nucleus (MGN; **A, B**) and lateral geniculate nucleus (LGN; **C, D**) seeds in healthy controls (HC; **A, C**) and people with schizophrenia (PSZ, **B, D**), from the RS+TL sample (55 PSZ, 46 HC; total N = 101). **A)** 25,166 significant grayordinates (2,971 positive, 22,195 negative). **B)** 23,177 significant grayordinates (2,012 positive, 21,165 negative). **C)** 27,497 significant grayordinates (4,417 positive, 23,080 negative). **D)** 21,820 significant grayordinates (5,312 positive, 16,508 negative). **E)** Whole-cortex t-statistic maps for group differences in LGN seed RSFC in the RS+TL sample, assessed using a design matrix with regressors for diagnosis, age, biological sex, and handedness. 1,824 significant grayordinates. Significance was assessed in Permutation Analysis of Linear Models, using 10,000 permutations or sign-flips and threshold-free cluster enhancement. P-values were corrected for family-wise error rate and t-statistic maps were thresholded at α = 0.05.

LGN was significantly connected in both groups to primary and secondary VC, posterior cingulate and retrosplenial cortex, and posterior PHA. Anticorrelations were observed with association AC, posterior parietal cortex, temporal-parietal-occipital junction, premotor cortex, motor cortex, and somatomotor cortex; in HC, additional anticorrelations were observed in left STG and tertiary VC.

Note that the use of global signal as a nuisance parameter when calculating seed correlations, while powerful for mitigating motion artifacts (71–74) that can especially confound studies of psychiatric populations (71, 74–81), effectively mean-centers seed correlation images such that all seed correlations across the brain approximately sum to zero. As a result, negative correlations in analyses controlling for global signal represent reduced connectivity relative to the whole-brain average, rather than truly observed anti-correlations (82), and should be interpreted as such.

#### Group Differences

No significant differences in whole-cortex MGN seed connectivity were observed between PSZ and HC. Whole-cortex LGN seed connectivity was significantly increased in PSZ relative to HC in bilateral somatosensory cortex, motor cortex, and tertiary VC, as well as right secondary VC **(Figure 2E).**

### Whole Thalamus Seed Connectivity

Regions showing significant seed connectivity to whole thalamus in each group are shown in **Figure 3A-B**. In both groups, thalamus was significantly connected to bilateral primary and secondary AC, primary VC, posterior cingulate and retrosplenial cortex, anterior cingulate cortex, and right anterior/middle insula, left ventral frontal opercular cortex, and right PHA. Anticorrelations were observed in bilateral motor, premotor and somatosensory cortex, secondary and tertiary VC, ventral visual cortical areas, large portions of superior and middle temporal gyri and sulci (including association AC), Wernicke’s area, temporal-parietal-occipital junction, posterior parietal cortex, and lateral PFC. PSZ additionally showed positive correlations with bilateral anterior and posterior insula, and HC anticorrelations with left granular insula.

**Figure 3.**
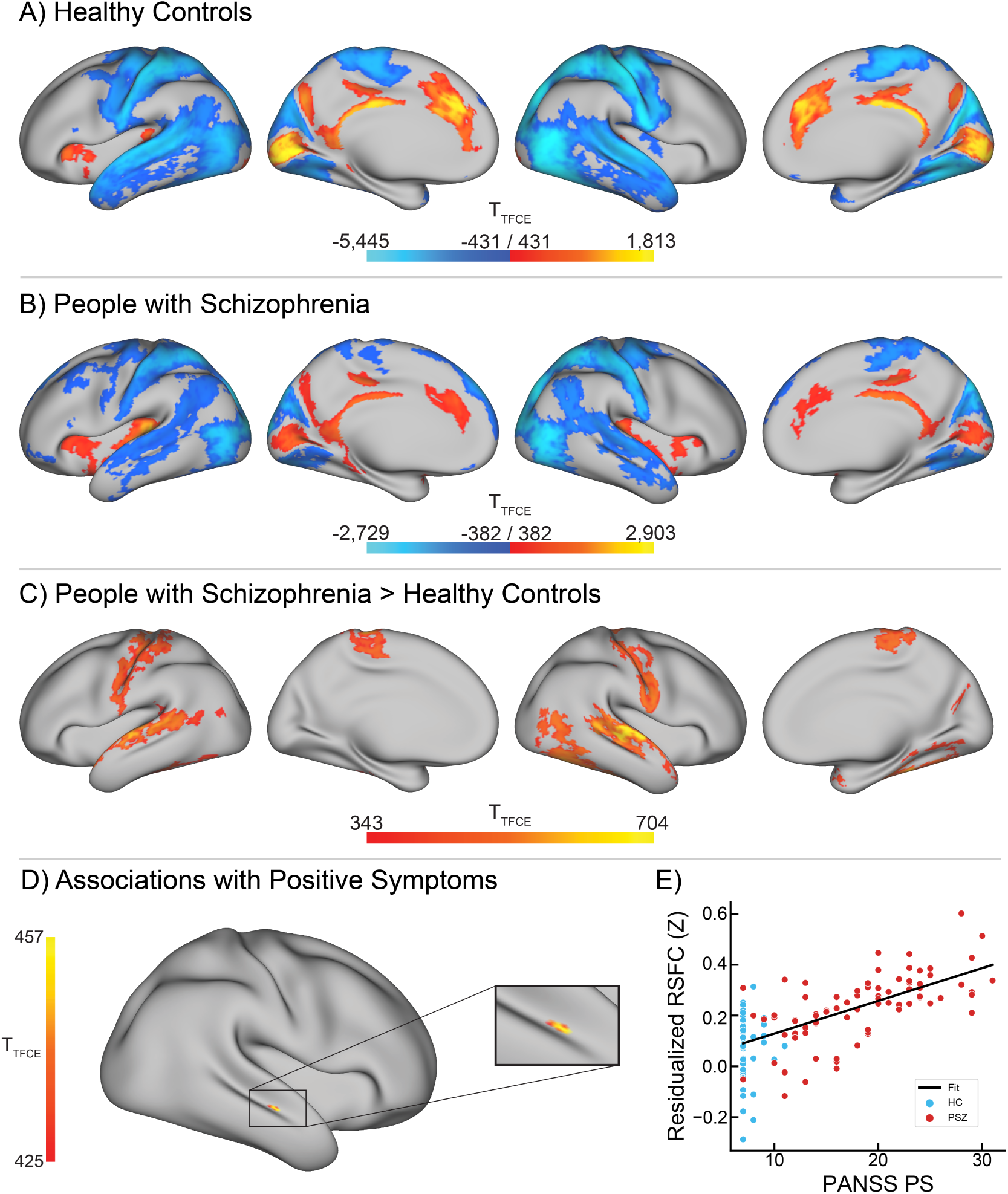
Panels **A-B** show whole-cortex t-statistic maps for grayordinates exhibiting significant resting-state functional connectivity (RSFC) from whole thalamus seeds in healthy controls (HC; **A**) and people with schizophrenia (PSZ, **B**), from the RS-only sample (82 PSZ, 55 HC; total N = 137). **A)** 28,971 significant grayordinates (4,095 positive, 24,876 negative). **B)** 23,347 significant grayordinates (4,551 positive, 18,796 negative). Panels **C-F** show t-statistic maps for whole thalamus seed resting-state functional connectivity (RSFC) group differences and associations with positive symptom severity RS-only sample. **C)** Whole thalamus seed RSFC group differences between people with schizophrenia (PSZ; n = 82), and healthy controls (HC; n = 55), controlling for age, biological sex, and handedness. 6,547 significant grayordinates. Total N = 137. **D)** Whole thalamus seed RSFC associations with PANSS Positive subscale scores, controlling for age, biological sex, handedness, and schizophrenia diagnosis, as well and PANSS negative and general subscores. PANSS scores were available for 78 PSZ and 51 HC (total N = 129). 9 significant grayordinates. **E)** Whole thalamus seed connectivity extracted from the significant grayordinates visualized in **D**, with thalamic RSFC residualized to covariates, displaying the identified relationship between thalamic seed connectivity and positive symptom severity. Significance was assessed grayordinate-wise in whole cortex using cortical surface GIFTIs in Permutation Analysis of Linear Models in whole cortex, using 10,000 permutations and threshold-free cluster enhancement. P-values were corrected for family-wise error rate and t-statistic maps were thresholded at α = 0.05.

#### Group Differences

Whole thalamus hyperconnectivity was observed in PSZ relative to HC in bilateral secondary and association AC (particularly STS and STG), somatosensory, premotor, and motor cortex, secondary and higher-order VC, PHA, and temporal-parietal-occipital junction **(Figure 3C**; unthresholded t-statistic maps shown in **Figure S5).**

#### Positive Symptom Associations

Whole thalamus hyperconnectivity was positively associated with positive symptom severity in right STS **(Figure 3D-E).**

## Discussion

The primary aim of this study was to translate findings of impaired auditory thalamocortical connectivity between medial geniculate nucleus (MGN) and primary auditory cortex (AC) in an animal model of 22q11DS (19, 30) to unmedicated PSZ, and reconcile these results with past findings of thalamocortical hyperconnectivity with sensory (and motor) cortex in past RSFC studies of PSZ (8, 9, 12, 14, 21). Contrary to our hypothesis, we found no group differences in connectivity between MGN and any region of AC, including transverse temporal gyrus (primary AC), as might have been expected for a direct translation of impairment in excitatory post-synaptic currents found in 22q11DS mice to humans with schizophrenia. However, we found that positive symptom severity was associated with *hyper*connectivity (but not hypoconnectivity) between MGN and bilateral superior temporal gyrus (STG; a region of association AC) **(Figure 1A-C).** This relationship was specific to positive symptoms (as analyses were performed while controlling for the severity of positive and negative symptoms, as well as schizophrenia diagnosis), and specific to connectivity from MGN **(Tables S1-S2).** This may indicate that hyperconnectivity in auditory cortico-basal ganglia-thalamo-cortical (CBGTC) circuits is specific to the presence and severity of positive symptoms, especially given the relative predominance of hallucinations in the auditory domain (41, 42). Whole thalamic hyperconnectivity was also positively associated with positive symptom severity in a region of right superior temporal sulcus **(Figure 3E-F);** this region was rather small, and likely reflects influence from auditory processing components of thalamocortical communication, as auditory processing regions are estimated to comprise about 8% of human cortex (83).

Conversely, connectivity from lateral geniculate nucleus (LGN) was significantly greater in PSZ relative to HC in secondary and higher-order areas of VC bilaterally **(Figure 1D-F);** this relationship was further found to be specific to connectivity from LGN **(Tables S4-S5).** In contrast to MGN-AC connectivity, no associations were found between LGN-VC connectivity and positive symptom severity. Whole-cortex follow-up analyses additionally revealed LGN hyperconnectivity with bilateral motor and somatosensory cortex **(Figure 2E)**. Similarly, whole thalamus seed connectivity was similarly increased in PSZ in large portions of bilateral secondary and association AC and VC and in somatosensory and motor cortical regions **(Figure 3C)**. While the whole-cortex thalamic seed connectivity analysis was likely more powered than using LGN (given the larger size and thereby improved ability to acquire signal in the whole thalamus ROI relative to the smaller and noisier geniculi), the broad similarity in group differences found using these seeds may indicate that widespread hyperconnectivity in secondary and higher-order thalamocortical sensory processing pathways could at least in part underlie consistent findings of thalamic hyperconnectivity to sensorimotor cortical regions in schizophrenia, especially given the substantial portion of the human cortex (estimated to be approximately 27%) that is predominately visual in function (83).

Notably, we did not replicate findings of thalamic hypoconnectivity with frontal cortex (7–10, 14, 16, 20, 84), in association with either schizophrenia diagnosis or positive symptom severity. This may be due to our fully unmedicated PSZ sample, either because unmedicated PSZ do not exhibit decreased thalamo-cortical connectivity with frontal cortex, or because our sample is higher functioning at baseline (i.e., without medication) than medicated PSZ samples. This result could potentially also be due to an insufficient sample size to detect this effect, although our findings are broadly in line with a prior study of human participants with 22q11DS (12). Further, in our study, both PSZ and HC were required to test negative for the use of recreational psychoactive substances (including delta-9-tetrahydrocannabinol) prior to the acquisition of fMRI data; as a result, the PSZ sample reported here could exhibit reduced hypofrontality (85, 86) relative to one that is more naturalistic. In unthresholded t-statistic maps of thalamic seed connectivity with whole cortex, we did observe trends towards hypoconnectivity in frontal cortex that did not reach the level of significance **(Figure S5).**

## Caveats and Limitations

The PSZ sample studied here is heterogenous in symptom severity **(Figure S1).** This may have provided power to detect symptom associations with MGN-AC hyperconnectivity as a primary analysis (as opposed to a secondary analysis using grayordinates extracted from another statistical test performed on the same data). However, if MGN-AC hyperconnectivity is truly more reflective of positive symptom severity than of overall diagnostic criteria for schizophrenia, the heterogeneity of the PSZ sample would similarly have reduced power for detecting group differences between PSZ and HC. Exploring MGN-AC hyperconnectivity as a transdiagnostic marker of positive symptom severity, however, would require future work involving the recruitment and study of participants across a range of disorders, in tandem with the use of the sensory TL task used here or other localization techniques.

It is plausible, however, that a primary impairment leading to pathologically reduced auditory thalamocortical connectivity is present in schizophrenia generally, and perhaps in relation to auditory hallucinations specifically, that was not detected in this study. For instance, while thalamocortical MGN-AC connectivity was impaired in the 22q11DS mouse, corticothalamic and cortico-cortico transmission auditory transmission were not (30). It is possible that substantial influence from other such pathways, which include polysynaptic connections that can impact RSFC (21), complicates the detection of thalamocortical connectivity deficits, which are additionally only detected bidirectionally using RSFC fMRI. It is important to note that our medial geniculate nuclei used here were whole MGN, while MGN-AC corticothalamic pathways found to be disrupted in the 22q11DS mouse were lemniscal, originating specifically in the ventral subnucleus of the MGN, and specifically terminating in primary AC (30). In contrast, signals propagating through non-lemniscal auditory circuits (87) may substantially contribute to measured MGN-AC RSFC. Further, primary thalamocortical auditory connectivity deficits may normalize (e.g., through compensatory mechanisms) over the course of brain development, and thus become unobservable, prior to the onset of psychosis.

## Conclusion

In this study of unmedicated PSZ and HC, we found that hyperconnectivity between MGN and AC is a specific correlate of positive symptom severity, though not of schizophrenia generally. Conversely, we found that connectivity between LGN and VC, as well as connectivity between whole thalamus and sensorimotor cortex, was increased in PSZ relative to HC, but not associated with positive symptom severity specifically. Overall, these results expand upon past findings from RSFC studies of PSZ and people with 22q11DS of thalamocortical hyperconnectivity to sensorimotor regions (1, 2, 4, 6–9, 11, 14, 15, 88) and establish that hyperconnectivity of MGN-AC circuits may be a marker of positive symptom severity, specifically, rather than of schizophrenia or risk for psychosis more generally.

## Data and Code Availability

The sensory Thalamic Localizer task Presentation code is available from GitHub at https://github.com/CNaP-Lab/Sensory-Thalamic-Localizer, as well as the Neurobehavioral Systems Archives of Neurobehavioral Experiments and Stimuli at http://www.neurobs.com/ex_files/expt_view?id=302. The MATLAB task analysis code for producing MGN and LGN fROIs from BOLD fMRI Thalamic Localizer task data and Presentation task logs is available from GitHub at https://github.com/CNaP-Lab/Sensory-Thalamic-Localizer. All software associated with this manuscript is released under the GNU General Public License version 3. Data acquired from human participants used in the analyses detailed in this manuscript are available upon request from the corresponding author through a formal data sharing agreement.

## Author Contributions

**Conceptualization:** G.H., A.A-D., and J.X.V.S.; **Data Curation:** J.C.W., P.N.T., R.B.G., Z.J.Z., E.B.S-F., N.K.H., S.K.A., D.T.P., N.O., K.B., M.S., J.J.W., G.P., G.H., A.A-D., and J.X.V.S.; **Formal Analysis:** J.C.W., P.N.T., and J.X.V.S.; **Funding Acquisition:** J.C.W., R.B.G., M.S., G.H., A.A-D., and J.X.V.S.; **Investigation:** J.C.W., P.N.T., R.B.G., Z.J.Z., E.B.S-F., N.K.H., S.K.A., D.T.P., N.O., K.B., M.S., J.J.W., G.P., G.H., A.A-D., and J.X.V.S.; **Methodology:** J.C.W., P.N.T., R.B.G., G.P., G.H., A.A-D., and J.X.V.S.; **Project Administration:** J.C.W., R.B.G., N.K.H., G.H., A.A-D., and J.X.V.S.; **Resources:** G.H., A.A-D., and J.X.V.S.; **Software:** J.C.W., P.N.T., S.K.A., and J.X.V.S.; **Supervision:** R.B.G., N.K.H., M.S., J.J.W., G.P., G.H., A.A-D., and J.X.V.S.; **Validation:** J.C.W., P.N.T., Z.J.Z., S.K.A, G.H., and J.X.V.S.; **Visualization:** J.C.W., P.N.T., D.T.P., and J.X.V.S.; **Writing – Original Draft:** J.C.W.; **Writing – Review & Editing:** J.C.W., P.N.T., G.P., G.H., A.A-D., and J.X.V.S.

## Data Availability

http://www.neurobs.com/ex_files/expt_view?id=302

## Acknowledgements

The authors would like to thank Jaeyop Jeong, Sam R. Luceno, Srineil Nizambad, and Yash Patel for contributions to fMRI data preprocessing, Isabella Rosario for assistance with data organization, and Tram N. B. Nguyen, Dr. Benjamin A. Ely, Dr. Aprajita Mohanty, and Dr. Christine DeLorenzo for helpful discussions. Computing resources and technical assistance were provided by Stony Brook Medicine Research Computing, with substantial support from Allen Zawada and James Xikis. Access to and technical support for the high-performance SeaWulf computing system was provided by Stony Brook Research Computing and Cyberinfrastructure and the Institute for Advanced Computational Science at Stony Brook University, with notable support from Fırat Coşkun, Daniel Wood, and David Carlson.

## Abbreviations

22q11DS: 22q11.2 microdeletion syndrome
AC: auditory cortex
BOLD: blood-oxygen-level dependent
CBGTC: cortico-basal ganglia-thalamo-cortical
CDSS: Calgary Depression Scale for Schizophrenia
CIFTI: Connectivity Informatics Technology Initiative
CSF: cerebro-spinal fluid
DSM-5: Diagnostic and Statistical Manual of Mental Disorders, Fifth Edition
DSM-IV-TR: Diagnostic and Statistical Manual of Mental Disorders, Fourth Edition, Text Revision
EHI: Edinburgh Handedness Inventory
FHS: Family History Screen
fMRI: functional magnetic resonance imaging
fROIs: functionally defined regions of interest
GS: General Psychopathology Scale
HC: healthy control
LGN: lateral geniculate nucleus
MGN: medial geniculate nucleus
MPs: motion parameters
MNI152NLin6: Montreal Neurological Institute 152 non-linear 6th-generation space
MSNs: medium spiny neurons
NIfTI: Neuroimaging Informatics Technology Initiative
NS: Negative Scale
NYSPI: New York State Psychiatric Institute
PANSS: Positive and Negative Syndrome Scale
PS: Positive Scale
PFC: prefrontal cortex
PSYRATS: Psychotic Symptom Rating Scales
PSZ: people with schizophrenia
ROIs: regions of interest
RS: resting-state
RSFC: resting-state functional connectivity
SANS: Scale for the Assessment of Negative Symptoms
SBU: Stony Brook University
SCID-5-RV: Structured Clinical Interview for DSM-5, Research Version
SCID-I: Structured Clinical Interview for DSM-IV Axis I Disorders
SES: socioeconomic status
T1w: T1-weighted
TL: Thalamic Localizer
VC: visual cortex
WM: white matter

