## Supplementary Material for "Auditory and Visual Thalamocortical Connectivity Alterations in Unmedicated People with Schizophrenia: An Individualized Sensory Thalamic Localization and Resting-State Functional Connectivity Study"

**Affiliations**

**Address:** 101 Nicolls Rd., Health Sciences Center T10-087J, Stony Brook, NY 11794

Short/running title: Auditory and Visual Thalamocortical Connectivity in Unmedicated People with Schizophrenia

All Participants 4
Healthy Control Participants (HC) 4
People With Schizophrenia (PSZ) 5

### Abbreviations

AC, auditory cortex;

BOLD, blood-oxygen-level dependent;

CSF, cerebro-spinal fluid;

DSM-5, Diagnostic and Statistical Manual of Mental Disorders, Fifth Edition;

DSM-IV-TR, Diagnostic and Statistical Manual of Mental Disorders, Fourth Edition, Text Revision;

EHI, Edinburgh Handedness Inventory;

fMRI, functional magnetic resonance imaging;

fROI, functionally defined region of interest;

HC, healthy control;

HRF, hemodynamic response function;

LGN, lateral geniculate nucleus;

LPF-FD, low-pass-filtered framewise displacement;

MacCAT-CR, MacArthur Competence Assessment Tool for Clinical Research;

MCOT, Multiband Censoring Optimization Tool;

MGN, medial geniculate nucleus;

MPs, motion parameters;

MR, magnetic resonance;

MRI, magnetic resonance imaging;

NIfTI, Neuroimaging Informatics Technology Initiative;

NYSPI, New York State Psychiatric Institute;

PANSS, Positive and Negative Syndrome Scale;

PSZ, people with schizophrenia;

ROIs, regions of interest;

RS, resting-state;

SBU, Stony Brook University;

SCID-5-RV, Structured Clinical Interview for DSM-5, Research Version;

SCID-I, Structured Clinical Interview for DSM-IV Axis I Disorders;

TL, Thalamic Localizer;

VC, visual cortex;

WM, white matter.

### Supplementary Materials and Methods

#### *Inclusion and Exclusion Criteria*

##### All Participants

Participants were recruited with advertisements, outpatient services at each institution, physician referrals, and via word-of-mouth. All individuals provided written consent prior to their participation and were free of psychoactive substances on the day of fMRI scanning as determined by a negative urine toxicology. PSZ were assessed for capacity to give informed consent using the MacArthur Competence Assessment Tool for Clinical Research (MacCAT-CR; 1) prior to obtaining informed consent. PSZ were either antipsychotic drug-naïve or antipsychotic drug-free for at least three weeks for reasons unrelated to this study prior to recruitment into this study.

All participants were free of any major neurological disorders, current substance use disorders, hearing impairment, and psychiatric disorders other than schizophrenia, schizophreniform disorder, or schizoaffective disorder. Psychiatric diagnoses at the New York State Psychiatric Institute (NYSPI) were established according to the Diagnostic and Statistical Manual of Mental Disorders, Fourth Edition, Text Revision (DSM-IV-TR; 2), using the Structured Clinical Interview for the DSM-IV Axis I Disorders (SCID-I; 3). Psychiatric diagnoses at Stony Brook University (SBU) were established according to the Diagnostic and Statistical Manual of Mental Disorders, Fifth Edition (DSM-5; 4), Structured Clinical Interview for DSM-5, Research Version (SCID-5-RV; 5).

All participants completed a magnetic resonance imaging (MRI) clearance form prior to any magnetic resonance (MR) scanning procedures to assess for the potential presence of metallic implants and past experiences with metal that could remain in the body. Participants were also administered a urine drug toxicology test before MR scanning procedures; in the event of a positive result for psychoactive drug use on the day of a scan, MR scanning was aborted and potentially rescheduled at the discretion of participants and study personnel, provided the participant did not meet criteria for a substance use disorder. Biologically female participants were administered a urine pregnancy screening test before MR scanning; in the event of a positive pregnancy test, the MR scan session would be aborted, and the participant would be excluded from further participation in the study.

##### Healthy Control Participants (HC)

##### *New York State Psychiatric Institute (NYSPI)*

At New York State Psychiatric Institute (NYSPI), inclusion criteria for healthy control (HC) participants were: 1) ages 18-55, 2) absence of any current DSM-IV-TR (2) Axis-I psychiatric disorder, and 3) fluent in spoken English.

NYSPI HC exclusion criteria were: 1) current substance use disorders; 2) current major affective episode; 3) history of neurological disorders, including head trauma with loss of consciousness, epilepsy, or other clinically significant disorders of the central nervous system, intellectual disability, hearing impairment, or unstable severe medical conditions;

4) claustrophobia; 5) metal implants or paramagnetic objects contained within the body; and 6) current pregnancy.

***Stony Brook University (SBU)***

At Stony Brook University (SBU), HC inclusion criteria were: 1) age 18-65 years old; 2) negative urine toxicology; 3) hearing threshold within normal limit (up to 25 decibels Hearing Level from 250 Hz through 2 kHz, bilaterally); 4) speech recognition for monosyllabic words equal to or better than 80% correct.

Exclusion criteria at SBU for HC were: 1) current or past, or family history of, psychiatric illness, including alcohol or substance abuse, except nicotine use disorder, per the DSM-5 (4); 2) clinically significant medical or neurological illness; 3) pregnancy or lactation; 4) lack of effective birth control (defined as use of either an intrauterine device, birth control pills, long-acting birth control injections, or both condoms and spermicide); 5) presence of metallic objects in the body; 6) current, past, or anticipated exposure to radiation in the workplaces, or participation in nuclear medicine procedures elsewhere; 7) current aural or neurological disease; 8) more than one risk factor for coronary disease (smoking, hyperlipidemia, or sedentary lifestyle); 9) insulin-dependent diabetes; 10) history of significant cardiovascular illness, determined using a cardiac screening questionnaire and a screening electrocardiogram; 11) stage 2 hypertension (blood pressure greater than 140 mm Hg systolic or 90 mm Hg diastolic); 12) clinically significant brain abnormalities; 13) prisoners; 14) severe claustrophobia; and 15) any previous severe adverse reaction to amphetamine.

**People With Schizophrenia (PSZ)**

All people with schizophrenia (PSZ) were either antipsychotic-free or antipsychotic-naïve for reasons unrelated to the study prior to their participation in the study. PSZ were considered to be antipsychotic medication-free if they had no exposure to oral antipsychotic medication for at least 3 weeks, and no exposure to intramuscular antipsychotic medications for at least 6 months, prior to their participation in this study. PSZ were additionally considered to be antipsychotic medication-naïve if they had fewer than 2 weeks of cumulative lifetime exposure to antipsychotic medications.

***New York State Psychiatric Institute (NYSPI)***

Inclusion criteria for NYSPI PSZ were: 1) diagnosis of schizophrenia or schizoaffective disorder, per the DSM-IV-TR; 2) antipsychotic medication-free or antipsychotic-naïve (defined above); 3) age 18-55 years old, 4) fluent in spoken English; and 5) capacity to give informed consent, as determined using the MacArthur Competence Assessment Tool for Clinical Research (MacCAT-CR; 1).

Exclusion criteria for NYSPI PSZ were: 1) current substance use disorders, 2) a current major affective episode, 3) history of neurological disorders, including head trauma with loss of consciousness, epilepsy, or other clinically significant disorders of the central nervous system, intellectual disability, hearing impairment, or unstable severe medical conditions, 4) claustrophobia; 5) metal implants or paramagnetic objects contained within the body; and 6) current pregnancy.

***Stony Brook University (SBU)***

Inclusion criteria for SBU PSZ were: 1) age 18-65 years old; 2) negative urine toxicology; 3) hearing threshold within normal limit (up to 25 decibels Hearing Level from 250 Hz through 2 kHz, bilaterally); 4) speech recognition for monosyllabic words equal to or better than 80% correct; 5) primary diagnosis of schizophrenia, schizophreniform disorder (if the diagnosis of schizophrenia was confirmed after six months), or schizoaffective disorder as per DSM-5 (4), which requires that psychosis is not attributable to other causes (general medical condition, substance-induced, dementia, etc.); 6) antipsychotic medication-free or antipsychotic-naïve (defined above), and 7) capacity to give informed consent, as determined using the MacArthur Competence Assessment Tool for Clinical Research (MacCAT-CR; 1).

Exclusion criteria for SBU PSZ were: 1) Major psychiatric disorder other than schizophrenia, schizophreniform, or schizoaffective disorder; 2) clinically significant medical or neurological illness; 3) pregnancy or lactation; 4) lack of effective birth control (defined as use of either an intrauterine device, birth control pills, long-acting birth control injections, or both condoms and spermicide); 5) presence of metallic objects in the body; 6) current, past, or anticipated exposure to radiation in the workplaces, or participation in nuclear medicine procedures elsewhere; 7) current aural or neurological disease; 8) more than one risk factor for coronary disease (smoking, hyperlipidemia, or sedentary lifestyle); 9) insulin-dependent diabetes; 10) history of significant cardiovascular illness, determined using a cardiac screening questionnaire and a screening electrocardiogram; 11) stage 2 hypertension (blood pressure greater than 140 mm Hg systolic or 90 mm Hg diastolic); 12) clinically significant brain abnormalities; 13) prisoners; 14) severe claustrophobia; and 15) any previous severe adverse reaction to amphetamine, or previous refusal to accept treatment with antipsychotic medications after amphetamine administration; 15) current use of amphetamines, opiates, cocaine, sedative-hypnotics, and/or cannabis, mood stabilizers in the last 4 weeks, or fluoxetine or monoamine oxidase inhibitors in the last 6 weeks; 16) considered to be unlikely to be able antipsychotic medication-free for the duration of the study (e.g. history of aggressive or self-injurious behavior, agitation, significant subjective distress/discomfort or command hallucinations while off of antipsychotic medications); 16) currently experiencing command hallucinations or admitted to an inpatient psychiatric unit for clinical reasons. 17) previous history of treatment refractoriness to all antipsychotic medications, including clozapine; 18) explicitly or implicitly indicates that monetary gain is the primary reason for study participation; or 19) documented or suspected prior substance or alcohol abuse history.

***Demographic and Clinical Assessments***

Symptom severity was assessed for PSZ and HC using Positive and Negative Syndrome Scale (PANSS; 6), and for a subset of participants using the Psychotic Symptom Rating Scales (PSYRATS; 7), Scales for the Assessment of Negative Symptoms (SANS; 8, 9), and Calgary Depression Scale for Schizophrenia (CDSS; 10, 11). Handedness was assessed using the Edinburgh Handedness Inventory (EHI; 12), and SES using the Hollingshead Four Factor Index of Socioeconomic Status (13). Family psychiatric history was assessed using the Family History Screen (FHS; 14). At SBU, hearing thresholds were evaluated either using a GSI Automated Method for Testing Auditory Sensitivity (AMTAS) Pro (Grason-Stadler, Eden Prairie, Minnesota) automated audiometry system, or by a licensed audiologist or otolaryngologist, and speech recognition for monosyllabic words using the Northwestern University Auditory Test No. 6 (15).

***fMRI Task Procedures***

Full details of the sensory Thalamic Localizer (TL) task and its validation are available elsewhere (see 16, see 17). In summary, participants are presented with 16 trials of alternating pseudorandom auditory or visual stimulation. The task uses a clustered-sparse temporal acquisition paradigm (18-20), in which fMRI volumes are not acquired during the presentation of task stimuli, to mitigate the confounding effect of scanner noise on measured responses to task stimuli (18). Each trial is 12 s in duration, and consists of a silence period, 9 s of stimulation to achieve peak hemodynamic response in sensory cortex (21, 22), a second silence period (equal in duration to the first), and the clustered acquisition of BOLD fMRI volumes. Auditory stimuli were presented as 9 pseudorandomly ordered sets of 900 ms music segments and 100 ms of silence. Music segments were extracted from the song, *Transmission94 (Parts 1 & 2)* by *Bonobo,* and normalized by mean amplitude across segments. Visual stimulation consisted of a circular black-and-white checkerboard with a central fixation cross, alternating between black and white with a 7.5 Hz reversal frequency. Task stimuli were presented using a dedicated laptop using Presentation software (Version 22.1, Build 10.23.20, Neurobehavioral Systems, Inc., Berkeley, CA). Auditory stimulation was delivered using a SereneSound headset and transducer (Resonance Technology, Inc., Northridge, CA), and visual stimulation was presented using a projector with a 60 Hz refresh rate. The TL task Presentation code is publicly available under the terms of the GNU Public License version 3 from GitHub at https://github.com/CNaP-Lab/Sensory-Thalamic-Localizer.

First-level modeling was performed using smoothed TL task fMRI data in SPM12 (23-25), version 7771, in MATLAB R2018a (The MathWorks, Inc., Natick, MA), to obtain beta estimates for auditory and visual betas, using a design matrix that additionally included nuisance regressors for intercept, 6 MPs and their squares, white matter (WM) signal, and cerebro-spinal fluid (CSF) signal, as well as two additional regressors for each run to model the second and third volumes of each sparse acquisition cluster, to account for T1 relaxation effects (18, 19, 26). Regressors were not convolved with the hemodynamic response function (HRF), as clustered-sparse temporal acquisition paradigm precludes sufficiently sampling the HRF.

Auditory cortex (AC) and visual cortex (VC) fROI search regions were generated using the Wake Forest University PickAtlas Toolbox (27, 28), from Broadmann Areas (BAs) 41 and 42, and BAs 17 and 18, respectively, which were then 3D dilated by one voxel. An [Auditory – Visual] contrast image was generated and used to localize the largest contiguous cluster in the AC and VC of each hemisphere maximally responsive to auditory or visual stimulation, respectively. Average AC and VC time series were then calculated within and across hemispheres.

Search regions for MGN and LGN were generated by dilating ROIs obtained by expanding the FreeSurfer thalamic nuclei segmentation (29, 30) and masking out surrounding structures. Voxels within this search region were assessed for coactivation with AC and VC using partial correlations with nuisance regressors for WM, CSF, gray matter signal, local WM, and a regressor each for the second and third image of each cluster. This generates coactivation maps with AC and VC, which are then thresholded. Suprathreshold voxels showing coactivation with both AC and VC are removed to ensure modality specificity, and the largest contiguous cluster for left and right auditory and visual thalamus are retained as fROIs for left and right MGN and LGN, respectively. Resulting MGN and LGN fROIs were evaluated by multiple members of the study team (JCW, PNT, ZJZ, and JXVS) by overlaying them over each participant's high-resolution anatomical images and temporally averaged TL task BOLD echo planar images; fROIs that considered to be anatomically implausible resulted in the exclusion that participant's TL fROIs from further analysis

Full details of this procedure are available elsewhere (16). The TL task analysis software is publicly available at no cost under the terms of the GNU Public License version 3 from GitHub at https://github.com/CNaP-Lab/Sensory-Thalamic-Localizer.

During resting-state (RS) scans, participants were instructed to remain still and awake with their eyes open, focused ahead on a fixation cross. One of each participant's eyes was monitored using an eye tracking camera (eye tracking data not collected) and eye closure times were recorded to exclude times from resting-state functional connectivity analyses during which participants were likely to have been sleeping, due to the potentially confounding effects of sleep states (31).

***fMRI Pre- and Post-Processing***

RS and TL fMRI data were preprocessed using the Human Connectome Project Minimal Preprocessing Pipelines, version 4.2.0 (32). Volumetric images were obtained in Neuroimaging Informatics Technology Initiative (NIfTI) format in Montreal Neurological Institute 152 non-linear 6th-generation space (MNI152NLin6; 33). Surfaces were obtained in CIFTI (Connectivity Informatics Technology Initiative) format, with surface registration using cortical surface matching (MSMsulc; 34, 35).

Six translational and rotational motion parameters (MPs) were obtained from the HCP MPP. White matter (WM) and cerebro-spinal fluid (CSF) nuisance signals were calculated from unsmoothed volumetric data as the average signal in all compartment voxels remaining after an iterative erosion procedure (36), wherein masks were eroded up to 3 times, as long as at least two voxels would remain after one additional erosion; global signal was calculated as the average signal from all in-brain voxels. RS and TL fMRI data were smoothed using a 4 mm full-width-half-maximum Gaussian filter. Volumes acquired during periods of excess motion (36, 37) or signal fluctuation (37-39) were removed by volume censoring (36, 37, 40-42) using LPF-FD and GEV-DV censoring thresholds (42) determined using the Multiband Censoring Optimization Tool (42), as described in the ***Volume Censoring*** section, below**.**

RS data were mode-1000 intensity normalized (36, 37), mean-centered, linearly detrended, and band-pass filtered using a 0.009-0.08 Hz second-order zero-phase Butterworth filter. Censored data were replaced using linear interpolation before band-pass-filtering, and then were discarded. MPs were additionally 0.009-0.08 Hz band-pass filtered to produce filtered MPs (fMPs) for use as nuisance regressors when generating seed RSFC images. The first and last 22 seconds of each run were then discarded to remove discontinuity artifacts after temporal filtering (22.1 s / 26 volumes at NYSPI, and 22.4 s / 28 volumes at SBU, due to slightly different TRs).

***Volume Censoring***

Volume censoring was performed using using study-wide thresholds for head motion (36, 37) using low-pass-filtered framewise displacement (LPF-FD; 42) and thresholds for signal fluctuation (37-39) determined for each run using run-adaptive GEV-DV censoring (42). For analyses using data available from all participants with usable TL MGN and LGN functionally defined regions of interest (fROIs), thresholds from the Multiband Censoring Optimization Tool (MCOT; 42) for LPF-FD ($\Phi_F$) and GEV-DV ($d_G$) were 76.09 mm and 6.9 (arbitrary units), respectively. For analyses using data from all participants at least 2 runs of usable RS data, thresholds obtained were 98.15 mm and 8.9 (arbitrary units), respectively. Volumes acquired during eye closures lasting longer than 3 seconds were censored. Volumes acquired between censored eye closures were additionally themselves censored if the time separating censored eye closure periods was less than 30 seconds. Contiguous clusters of data shorter than 8 s in duration after censoring were removed as well; any resting-state runs with less than 1.5 minutes of remaining data were subsequently discarded. After censoring, participants with fewer than 2 runs of RS data, or fewer than 5 minutes of RS data total, were excluded from further analyses.

***Post-Hoc Specificity Analyses***

We sought to ensure that associations between positive symptom severity and MGN-AC connectivity were not better explained by relationships with connectivity from either LGN or MD (as nuclei involved in another sensory modality and higher-order cognition, respectively). Mean connectivity estimates from all 3 nuclei (MGN, LGN, and MD) were spatially averaged within the region of AC exhibiting these significant associations (“significant MGN-AC”; see **Figure 1B**) and entered as regressors into a linear model with PANSS Positive as the dependent variable and additional regressors for PANSS Negative, PANSS General, diagnosis, and demographic covariates (biological sex, age, and handedness). This was performed similarly for detected group differences in LGN-VC, extracting LGN, MGN, and MD connectivity estimates from the region of VC showing significant relationships between LGN connectivity and diagnosis (“significant

LGN-VC”; see **Figure 1E**); these were entered as regressors in a linear model with diagnosis as the dependent variable and additional regressors for demographic covariates.

To determine whether associations between MGN RSFC and positive symptom severity were specific to signal from MGN (rather than pan-thalamic), we then produced additional MGN seed connectivity images that included signal from MD and LGN as additional nuisance regressors (as described in **Seed Resting-State Functional Connectivity**). From these images, we extracted mean MGN connectivity within the significant MGN-AC region and entered it as the dependent variable in a linear model with regressors for PANSS Positive, PANSS Negative, PANSS General, diagnosis, and biological covariates. We tested the PANSS PS term to determine whether the extracted relationship was robust to controlling for signal from MD and LGN. This was repeated using a version of LGN seed connectivity that was calculated while additionally controlling for MGN and MD signals, except using a design matrix that included only terms for diagnosis and biological covariates.

We additionally tested whether controlling for MD and LGN signals produced a significant change in the relationship between MGN-AC connectivity and positive symptom severity in this region using a linear mixed effects model (43, 44). MGN-AC connectivity before and after controlling for MD/LGN signals was input as the dependent variable (repeated measures); the model included random effects of participant-specific intercepts, and fixed effects for biological covariates, diagnosis, PANSS Positive, PANSS Negative, and PANSS General, a “condition” term denoting whether connectivity estimates were controlling for MD/LGN signal, and a “condition” × PANSS Positive interaction term. The significance of the impact of controlling for MD and LGN signals on the detected relationship between MGN-AC connectivity and PANSS Positive was determined by testing the interaction term. This was also repeated using the LGN-VC connectivity (in place of MGN-AC connectivity), with identical random effects, and fixed effects for biological covariates, diagnosis, a “condition” term denoting whether connectivity estimates were controlling for MD/MGN signal, and a “condition” × diagnosis interaction term. The significance of the impact of controlling for MD and MGN signals on detected LGN-VC group differences was determined by testing the interaction term.

***Multiple Imputation***

Missing values for age, biological sex, handedness, race, ethnicity, and parental socioeconomic status, as well as the positive, negative, and general subscales from the Positive and Negative Syndrome Scale (PANSS; 6), were estimated with multiple imputation, separately for PSZ and HC, using SPSS Statistics (Version 29.0.0.0, Build 241, International Business Machines Corporation, Armonk, NY), as the aggregate median of 1,000 imputations. Each imputation was set to include a maximum of 100 Markov chain Monte Carlo iterations, using linear regression models for scale variables, and two-way interactions among categorical predictors. Singularity tolerance was $10^{-12}$. Maximum case draws and maximum parameter draws were set to 1,000. Imputed demographic data were used as covariates in the

ComBat site harmonization (45-48) procedure and as covariates in group-level analyses. Imputed PANSS assessment data were used only for ComBat site harmonization and were not used in primary group-level analyses exploring relationships with symptom severity.

### Supplementary Tables

**Table S1.** Regression model for positive symptom severity including regressors for resting-state functional connectivity from medial geniculate nucleus (MGN), lateral geniculate nucleus (LGN), and mediodorsal nucleus (MD), with the region of auditory cortex (AC) wherein MGN connectivity was found to be significantly associated with positive symptom severity.

| Effect | Estimate (*B*) | *SE* (*B*) | β | $t_{84}$ | p |
|---|---|---|---|---|---|
| Intercept | -1.056 | 2.041 | — | -0.5174 | 0.6062 |
| Diagnosis (Schizophrenia) | 3.449 | 1.142 | 0.2340 | 3.020 | $3.346\times10^{-3}$ |
| Age (Years) | 0.05522 | 0.03355 | 0.08004 | 1.646 | 0.1035 |
| Biological Sex (Male) | 0.6988 | 0.7686 | 0.04429 | 0.9092 | 0.3658 |
| Handedness (Right) | 0.4517 | 1.4932 | 0.01419 | 0.3025 | 0.7630 |
| PANSS Negative | -0.1383 | 0.08724 | -0.1035 | -1.585 | 0.1167 |
| PANSS General | 0.4432 | 0.05051 | 0.6972 | 8.774 | $1.694\times10^{-13}$ |
| MGN-AC Connectivity | 14.79 | 4.969 | 0.1789 | 2.976 | $3.819\times10^{-3}$ |
| LGN-AC Connectivity | 4.598 | 4.474 | 0.05974 | 1.0277 | 0.3070 |
| MD-AC Connectivity | 0.2043 | 3.406 | $3.931\times10^{-3}$ | 0.05997 | 0.9523 |

*Note.* N = 94, Error degrees of freedom = 84, R-squared = 0.81, Adjusted R-squared = 0.789, F-statistic versus constant model = 39.7, p-value = $1.30\times10^{-26}$. Model coefficients estimated using Huber robust regression. Symptom severity was assessed using the positive and negative syndrome scale (PANSS), positive, negative, and general symptom scales.
*Abbreviations*: PANSS: Positive and Negative Syndrome Scale; MGN: medial geniculate nucleus; LGN: lateral geniculate nucleus; MD: mediodorsal nucleus; AC: auditory cortex; *B*: unstandardized model coefficient; SE: standard error; β: standardized model coefficient; N: number of observations.

**Table S2.** Regression model for resting-state functional connectivity between medial geniculate nucleus (MGN) and the region of auditory cortex wherein MGN connectivity was found to be significantly associated with positive symptom severity, calculated while controlling for signals from mediodorsal and lateral geniculate thalamic nuclei.

| Effect | Estimate (*B*) | *SE* (*B*) | β | $t_{86}$ | p |
|---|---|---|---|---|---|
| Intercept | 0.03945 | 0.03579 | — | 1.102 | 0.2734 |
| Diagnosis (Schizophrenia) | -0.01039 | 0.02080 | -0.08083 | -0.4995 | 0.6187 |
| Age (Years) | $-1.065\times10^{-3}$ | $6.067\times10^{-4}$ | -0.1791 | -1.755 | 0.08285 |
| Biological Sex (Male) | .01489 | 0.01388 | 0.1096 | 1.073 | 0.2861 |
| Handedness (Right) | $7.233\times10^{-3}$ | 0.02688 | 0.02692 | 0.2691 | 0.7885 |
| PANSS Positive | $7.006\times10^{-3}$ | $1.774\times10^{-3}$ | 0.8044 | 3.948 | $1.604\times10^{-4}$ |
| PANSS Negative | $2.009\times10^{-3}$ | $1.591\times10^{-3}$ | 0.1726 | 1.263 | 0.2101 |
| PANSS General | $-4.565\times10^{-3}$ | $1.226\times10^{-3}$ | -0.8246 | -3.724 | $3.494\times10^{-4}$ |

*Note.* N = 94, Error degrees of freedom = 89, R-squared = 0.196, Adjusted R-squared = 0.131, F-statistic versus constant model = 3.0, p-value = $7.26\times10^{-3}$. Model coefficients estimated using Huber robust regression. Symptom severity was assessed using the positive and negative syndrome scale (PANSS), positive, negative, and general symptom scales.
*Abbreviations*: MGN: medial geniculate nucleus; *B*: unstandardized model coefficient; SE: standard error; β: standardized model coefficient; N: number of observations; PANSS: Positive and Negative Syndrome Scale.

**Table S3.** Linear mixed-effects model for resting-state functional connectivity between medial geniculate nucleus (MGN) and the region of auditory cortex wherein MGN connectivity was found to be significantly associated with positive symptom severity, calculated both with and without controlling for signals from lateral geniculate nucleus (LGN) and mediodorsal nucleus (MD).

| Random Effects | | *SD* | | | |
|---|---|---|---|---|---|
| Participant Intercept | | 0.06614 | | | |
| **Fixed Effects** | **Estimate (*B*)** | ***SE* (*B*)** | **β** | **$t_{178}$** | **p** |
| Intercept | $2.889\times10^{-3}$ | 0.04129 | — | 0.06997 | 0.9443 |
| Diagnosis (Schizophrenia) | -0.01403 | 0.02386 | -0.08833 | -0.588 | 0.5573 |
| Age (Years) | $-7.104\times10^{-4}$ | $6.958\times10^{-4}$ | -0.09671 | -1.021 | 0.3087 |
| Biological Sex (Male) | $6.134\times10^{-3}$ | 0.01591 | 0.03653 | 0.3855 | 0.7003 |
| Handedness (Right) | -0.02167 | 0.03083 | -0.06528 | -0.7028 | 0.4831 |
| PANSS Positive | 0.01037 | $2.055\times10^{-3}$ | 0.9632 | 5.044 | $1.118\times10^{-6}$ |
| PANSS Negative | $1.897\times10^{-3}$ | $1.824\times10^{-3}$ | 0.1319 | 1.040 | 0.2999 |
| PANSS General | $-5.006\times10^{-3}$ | $1.406\times10^{-3}$ | -0.7319 | -3.561 | $4.740\times10^{-4}$ |
| Condition: Controlling for MD and LGN Signals | 0.06269 | $8.846\times10^{-3}$ | 0.3970 | 7.086 | $3.099\times10^{-11}$ |
| PANSS Positive × Condition | $-2.459\times10^{-3}$ | $5.726\times10^{-4}$ | -0.2664 | -4.295 | $2.865\times10^{-5}$ |

*Note.* N = 94, AIC = - 461.51, BIC = -423.32, Log-Likelihood = 242.75, Deviance = -485.51. Model parameters estimated using restricted maximum likelihood estimation. “Controlling for MD and LGN Signals” fixed effects regressors indicate whether MGN seed connectivity was calculated while controlling for signals from mediodorsal nucleus (MD) and lateral geniculate nucleus (LGN).
*Abbreviations*: MGN: medial geniculate nucleus; LGN: lateral geniculate nucleus; MD: mediodorsal nucleus; VC: visual cortex; SD: standard deviation; *B*: unstandardized model coefficient; SE: standard error; β: standardized model coefficient; N: number of observations.

**Table S4.** Regression model for schizophrenia diagnosis including regressors for resting-state functional connectivity from lateral geniculate nucleus (LGN), medial geniculate nucleus (MGN), and mediodorsal nucleus (MD), with the region of visual cortex (VC) wherein LGN connectivity was found to be significantly associated with diagnosis.

| Effect | Estimate (*B*) | *SE* (*B*) | β | $t_{94}$ | p |
|---|---|---|---|---|---|
| Intercept | 0.5549 | 0.2333 | — | 2.379 | 0.01939 |
| Age (Years) | $6.566\times10^{-3}$ | $4.488\times10^{-3}$ | 0.1403 | 1.463 | 0.1468 |
| Biological Sex (Male) | 0.09546 | 0.1014 | 0.08918 | 0.9415 | 0.3489 |
| Handedness (Right) | -0.1744 | 0.2001 | -0.08073 | -0.8716 | 0.3857 |
| MGN-VC Connectivity | -0.5653 | 0.8726 | -0.0734 | -0.6478 | 0.5187 |
| LGN-VC Connectivity | 2.751 | 0.7087 | 0.5058 | 3.881 | $1.929\times10^{-4}$ |
| MD-VC Connectivity | 0.1629 | 0.6506 | 0.03662 | 0.2504 | 0.8028 |

*Note.* N = 101, Error degrees of freedom = 94, R-squared = 0.242, Adjusted R-squared = 0.194, F-statistic versus constant model = 5.02, p-value = $1.69\times10^{-4}$. Model coefficients estimated using Huber robust regression.
*Abbreviations*: LGN: lateral geniculate nucleus; MGN: medial geniculate nucleus; MD: mediodorsal nucleus; VC: visual cortex; *B*: unstandardized model coefficient; SE: standard error; β: standardized model coefficient; N: number of observations.

**Table S5.** Regression model for resting-state functional connectivity between lateral geniculate nucleus (LGN) and the region of visual cortex wherein LGN connectivity was found to be significantly associated with diagnosis (schizophrenia versus healthy control), calculated while controlling for signals from mediodorsal and medial geniculate thalamic nuclei.

| Effect | Estimate (*B*) | *SE* (*B*) | β | $t_{96}$ | p |
|---|---|---|---|---|---|
| Intercept | -0.04359 | 0.02872 | — | -1.518 | 0.1323 |
| Diagnosis (Schizophrenia) | 0.06255 | 0.01164 | 0.4477 | 5.375 | $5.359\times10^{-07}$ |
| Age (Years) | $-4.872\times10^{-4}$ | $5.531\times10^{-4}$ | -0.07448 | -0.8808 | 0.3806 |
| Biological Sex (Male) | -0.02749 | 0.01247 | -0.1838 | -2.204 | 0.02993 |
| Handedness (Right) | 0.02734 | 0.02535 | 0.09055 | 1.078 | 0.2837 |

*Note.* N = 101, Error degrees of freedom = 96, R-squared = 0.271, Adjusted R-squared = 0.241, F-statistic versus constant model = 8.92, p-value = $3.61\times10^{-6}$. Model coefficients estimated using Huber robust regression.
*Abbreviations*: LGN: lateral geniculate nucleus; *B*: unstandardized model coefficient; SE: standard error; β: standardized model coefficient; N: number of observations.

**Table S6.** Regression model for schizophrenia diagnosis including regressors for resting-state functional connectivity from lateral geniculate nucleus (LGN), medial geniculate nucleus (MGN), and mediodorsal nucleus (MD), with the region of visual cortex wherein LGN connectivity is significantly associated with diagnosis.

| Random Effects | | *SD* | | | |
|---|---|---|---|---|---|
| Participant Intercept | | 0.06649 | | | |
| **Fixed Effects** | **Estimate (*B*)** | ***SE* (*B*)** | **β** | **$t_{181}$** | **p** |
| Intercept | -0.05650 | 0.03476 | — | -1.626 | 0.1058 |
| Diagnosis (Schizophrenia) | 0.08739 | 0.01500 | 0.5153 | 5.827 | $2.538\times10^{-8}$ |
| Age (Years) | $-1.103\times10^{-3}$ | $6.796\times10^{-4}$ | -0.1407 | -1.623 | 0.1062 |
| Biological Sex (Male) | -0.04018 | 0.01529 | -0.2240 | -2.628 | $9.324\times10^{-3}$ |
| Handedness (Right) | 0.02261 | 0.03047 | 0.0638 | 0.7421 | 0.4590 |
| Condition: Controlling for MD and MGN Signals | 0.03654 | $5.929\times10^{-3}$ | 0.2167 | 6.163 | $4.527\times10^{-9}$ |
| Diagnosis (Schizophrenia) × Condition | -0.01823 | $7.971\times10^{-3}$ | -0.09671 | -2.287 | 0.02337 |

*Note.* N = 94, AIC = -514.1, BIC = -485.3, Log-Likelihood = 266.0, Deviance = -532.1. Model parameters estimated using restricted maximum likelihood estimation. “Controlling for MD and MGN” Signals fixed effects regressors indicate whether LGN seed connectivity was calculated while controlling for signals from mediodorsal nucleus (MD) and medial geniculate nucleus (MGN).
*Abbreviations*: LGN: lateral geniculate nucleus; MGN: medial geniculate nucleus; MD: mediodorsal nucleus; SD: standard deviation; *B*: unstandardized model coefficient; SE: standard error; β: standardized model coefficient; N: number of observations.

### Supplementary Figures

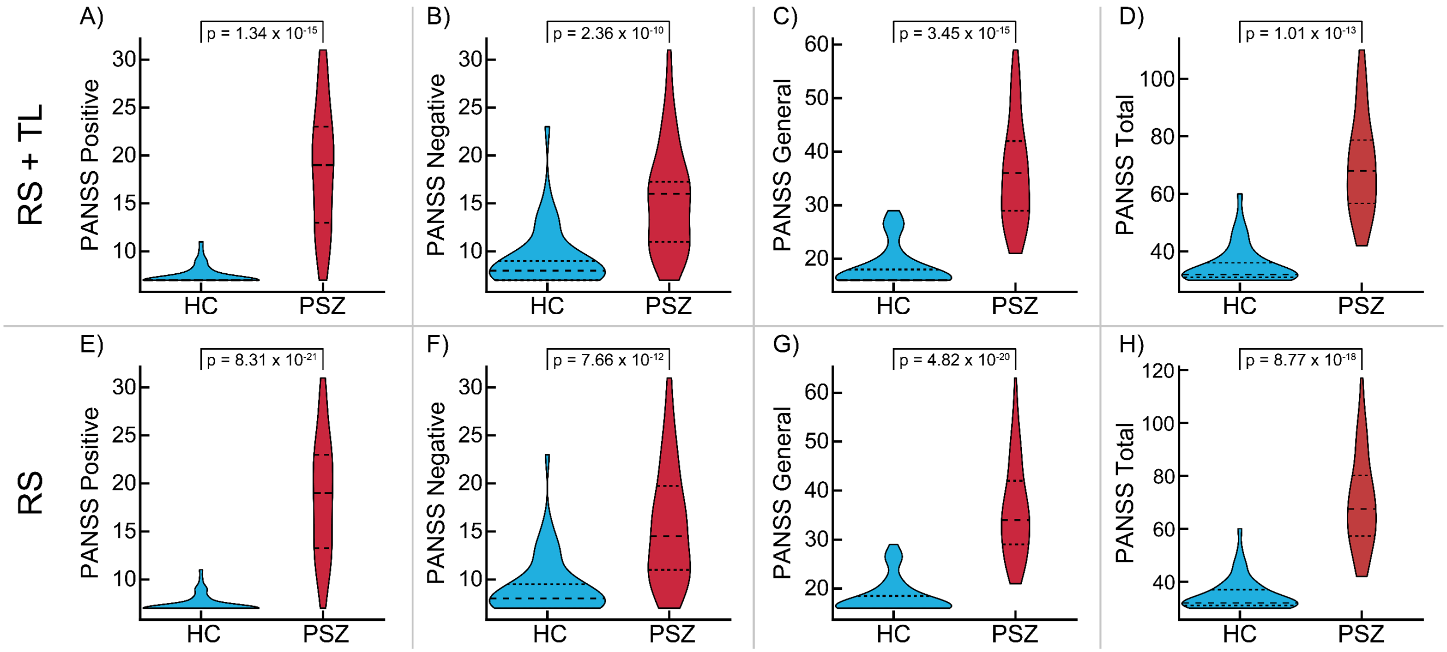

**Figure S1.** Positive and Negative Syndrome Scale (PANSS) Positive **(A, E)**, Negative **(B, F)**, and General **(C, G)** symptom subscale scores, and total across all subscales **(D, H)**, in healthy controls (HC, blue) and people with schizophrenia (PSZ, red). Scores are shown for the resting-state and thalamic localizer sample (RS+TL; top row), and the resting-state only sample (RS; bottom row). Violin inlays denote quartiles. P-values are shown for group differences determined using Mann-Whitney-Wilcoxon rank-sum tests.

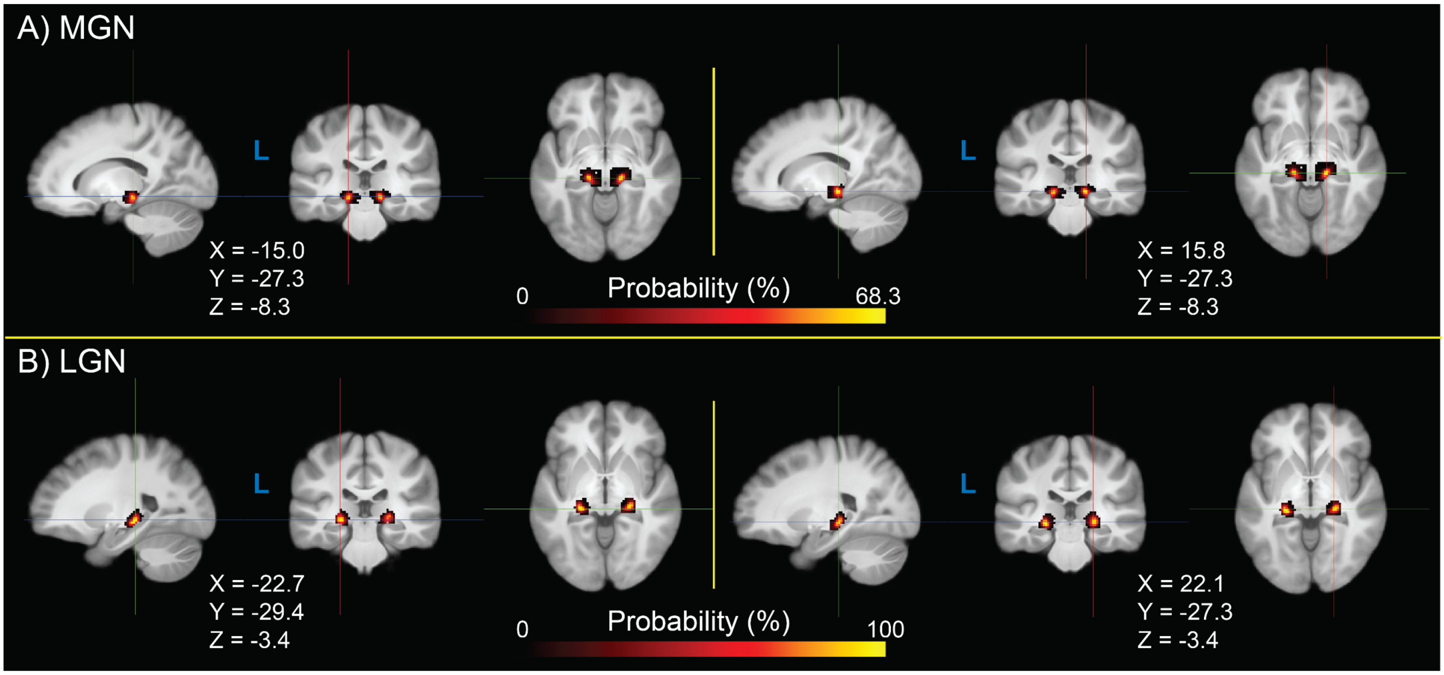

**Figure S2.** Probability density of bilateral medial geniculate nucleus (MGN; **A**) and lateral geniculate nucleus (LGN; **B**) regions of interest obtained from the thalamic localizer task across participants in the RS+TL sample (55 people with schizophrenia, 46 healthy controls; total N = 101), in Montreal Neurological Institute 152 non-linear 6th-generation (MNI152NLin6) space. Crosshair coordinates are shown for each view.

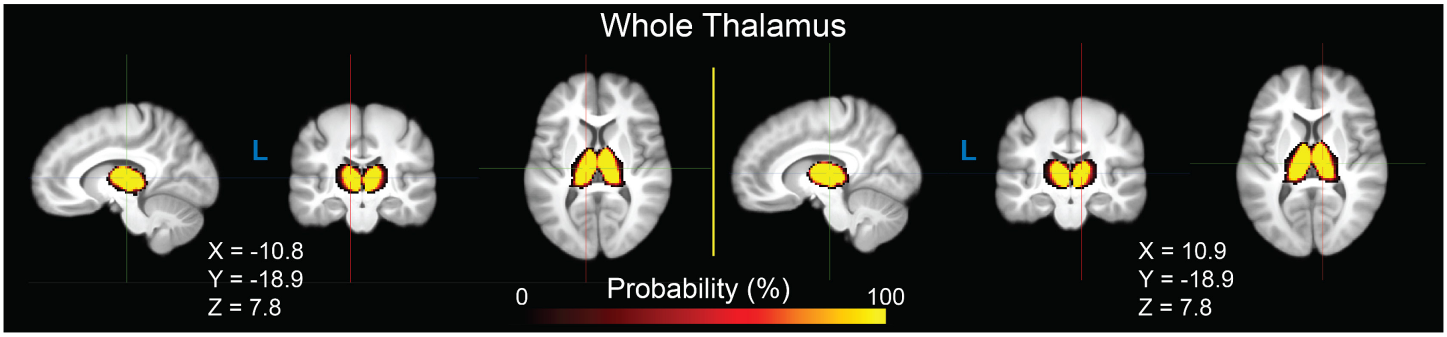

**Figure S3.** Probability density of bilateral whole thalamus regions of interest across all participants who completed resting-state fMRI (RS-only; 82 people with schizophrenia, 55 healthy controls; total N = 137), in Montreal Neurological Institute 152 non-linear 6th-generation (MNI152NLin6) space. Crosshair coordinates are shown for each view.

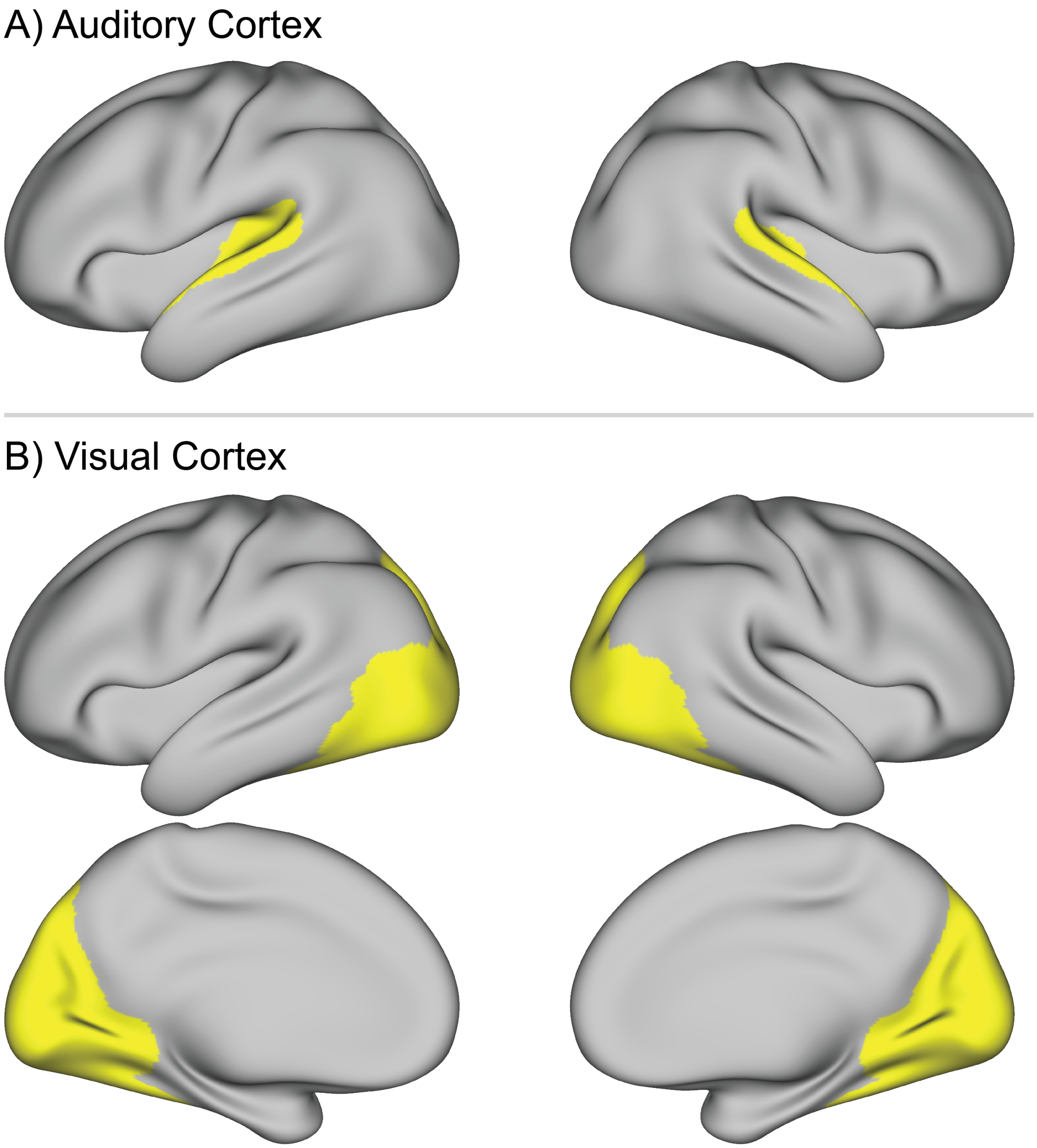

**Figure S4.** Bilateral auditory cortex **(A)** and visual cortex **(B)** regions of interest (ROIs) used in group-level seed resting-state functional connectivity permutation analyses, generated using the Cole-Anticevic Brain Network Partition of the Glasser cortical parcellation. The auditory cortex ROI was generated by selecting all parcels in the “auditory” network, dilating each by 4mm, and uniting them. The visual cortex ROI was generated by selecting all parcels in either the ”visual1” network or the “visual2” network, dilating by 4mm, and uniting them.

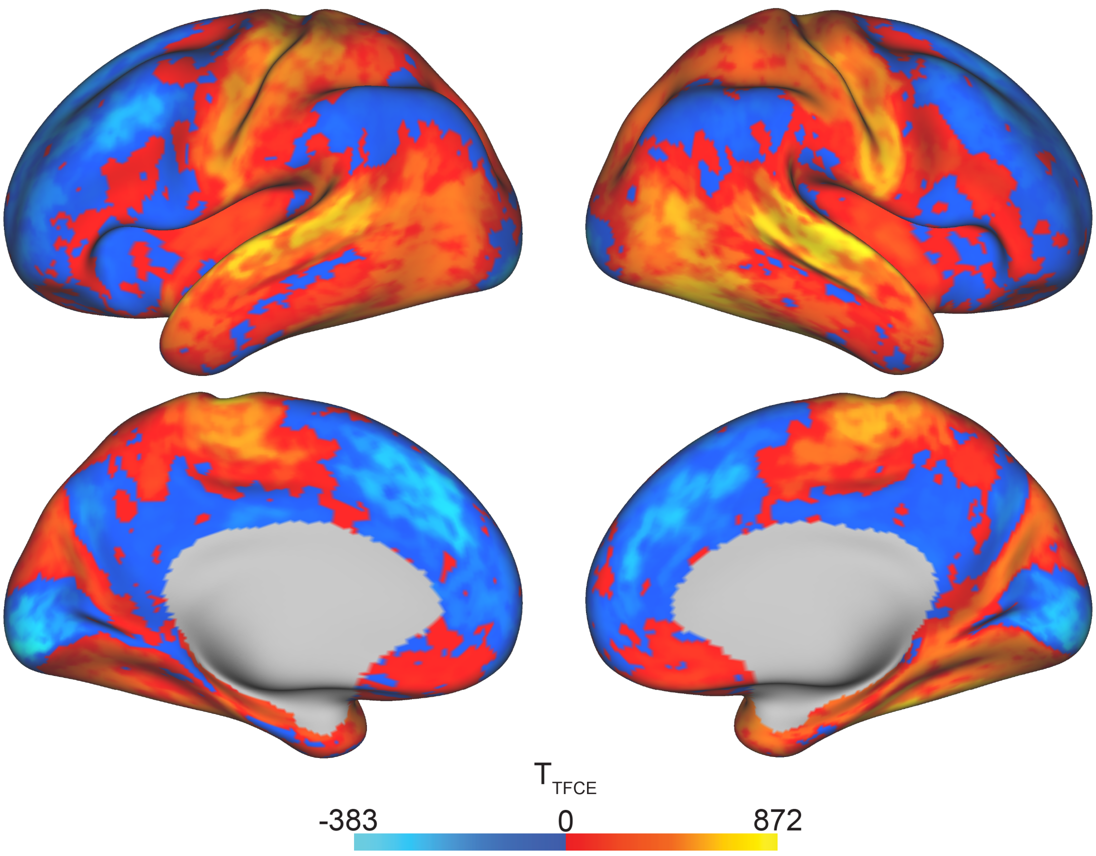

**Figure S5.** Unthresholded t-statistic maps for group differences in whole thalamus seed resting-state functional connectivity between people with schizophrenia (PSZ; n = 82) and healthy controls (HC; n = 55) in the RS-only case-control sample (total N = 137) T-statistics were calculated in whole cortex using Permutation Analysis of Linear Models, using 10,000 permutations and threshold-free cluster enhancement, using a design matrix including a schizophrenia diagnosis regressor and covariates for age, biological sex, and handedness.
